# State-by-State prediction of likely COVID-19 scenarios in the United States and assessment of the role of testing and control measures

**DOI:** 10.1101/2020.04.24.20078774

**Authors:** Zheng-Meng Zhai, Yong-Shang Long, Jie Kang, Yi-Lin Li, Lang Zeng, Li-Lei Han, Zhao-Hua Lin, Ying-Qi Zeng, Da-Yu Wu, Ming Tang, Di Xu, Zonghua Liu, Ying-Cheng Lai

**Author notes:** These authors contributed equally to this work (Zheng-Meng Zhai, Yong-Shang Long).

## Abstract

Due to the heterogeneity among the States in the US, predicting COVID-19 trends and quantitatively assessing the effects of government testing capability and control measures need to be done via a State-by-State approach. We develop a comprehensive model for COVID-19 incorporating time delays and population movements. With key parameter values determined by empirical data, the model enables the most likely epidemic scenarios to be predicted for each State, which are indicative of whether testing services and control measures are vigorous enough to contain the disease. We find that government control measures play a more important role than testing in suppressing the epidemic. The vast disparities in the epidemic trends among the States imply the need for long-term placement of control measures to fully contain COVID-19.

## INTRODUCTION

On about April 3, 2020, the United States registered the largest number of confirmed patients with the 2019 novel coronavirus (COVID-19) among all countries in the world: over 260,000 with the number of deaths exceeding 6,600. As of April 16, there had been over 641,000 cases and over 31,000 deaths. This development is rather astonishing, considering that there were only a few confirmed cases two months ago, when a massive outbreak in China had occurred. In fact, from the end of January, for various reasons the number of reported cases in the US increased quite slowly for a stretched period of time. The onset of an exponential increase in the US occurred in the middle of March, where it became apparent that COVID-19 began the phase of community spreading. In response, the White House issued a nationwide social-distancing order on March 16. Statewide stay-at-home or shelter-in-place orders were given by the governors of various States at different time (at the time of writing, there are still seven States that have not issued such an order). To quantitatively predict the effectiveness of the federal and State government measures to control COVID-19 spreading in the United States is utterly urgent. (Here we use “State” to denote a State in the US to distinguish it from a “state” as in an epidemic state.)

The United States differs from other countries in that the circumstances under which COVID-19 spreads vary dramatically among different States: not only are the levels of travel restriction orders dissimilar, but other factors affecting the disease spreading such as the population, medical resources, and social/political attitudes are also distinct among the States. A quantitative assessment of the effects of the control measures taken by the government to contain the COVID-19 pandemic thus needs to be carried out on a State-by-State basis. A complication is that each individual State is not a closed system: people move into and out of the State on a daily basis. This presents a tremendous challenge to modeling, as the existing data analyses and models for COVID-19 were mainly for the setting of a closed system [1–23] without considering the inbound and outbound population movements. [An introductory description of the recent work on data analysis and modeling of COVID-19 is presented in Supplementary Note (SN) 1.] In this paper, we meet this challenge by developing a coupled, dual-system spreading model. In particular, to predict the epidemic trend for any specific target State in the US, we treat the target system (A) as one under influences from another, much larger system (B) that represents all the other States. Because the size of B is much larger than that of A, in terms of the spreading dynamics, system B can be regarded as a closed system. From the standpoint of nonlinear physics, the influences of system B on system A can be viewed as a perturbation or background noise, while the effects of A on B can be neglected. The perturbation can be estimated based on the population of the target State and the empirical human movement data. The backbone of this unidirectionally coupled system is our recently developed, non-Markovian, five-state spreading model incorporating various time delays that are characteristic of COVID-19, which has been demonstrated to have the power to accurately predict the epidemic trends in China, South Korea, Italy, Iran, and the United Kingdom [24].

We select ten representative States in the US and aim to predict, for each State, how two key parameters affect the size of the final infected population and the epidemic duration: one characterizing the testing capability of the State, which essentially determines the fraction of undocumented infected population, and another quantifying the strength of the government actions. Using the daily number of confirmed cases up to March 29 to estimate the basic parameters, the model can generate a number of distinct epidemic trajectories into the future for each State. A comparison with the available data after March 29 enables us to pin down the specific scenario(s) for each State, attesting to the predictive power of the model. The predicted scenarios vary among the States and they are indicative of whether testing services and control measures are vigorous enough to fully contain the spreading of COVID-19. Systematic simulations reveal that, while sufficient testing can be beneficial, the control measures play a more important role in suppressing the epidemic, where strict government actions can reduce the epidemic duration and size in an exponential manner. An alarming result is that the length of the epidemic duration can differ by as much as one year among the States, posing a grave challenge to the government efforts in suppressing the pandemic and implying the continuous need for long-term and vigorous enforcement of the current control measures.

## RESULTS

We present prediction results from our generalized, dual coupled system, five-state model (described in Methods) for ten States: New York (the current epicenter), Washington (the previous epicenter), New Jersey, California, Michigan, Florida, Illinois, Massachusetts, Louisiana, and Arizona. (Source of data - Center for Systems Science and Engineering at Johns Hopkins University [25]). State-wise, up to March 29, the first nine States in this list had the top nine largest numbers of confirmed cases among the 50 States. The last State in the list, Arizona, is the home State of one of the co-authors. We use the daily number of confirmed cases up to March 29 to estimate the model parameters. For each State, we generate a number of epidemic scenarios and use the empirical data after March 29 to determine the most likely one(s). Detailed results for Arizona, Washington, and New York are described below, while the results for the remaining seven States are presented as SNs. In all the cases, each State is an open system with the influences of the rest of the country treated as a perturbation. Predictions of the epidemic trend of the entire country are also included as an SN. (Because the US is effectively a closed system, in this case the generalized coupling model is not necessary; instead, the single system, five-state model that we have developed recently [24] suffices.) For quantitatively assessing the effects of interstate travel on the epidemic through comparison, we also include results from the closed system model for two States: New York and Arizona.

In addition to predicting the most likely epidemic scenario(s) for each State, we seek to quantitatively assess the impacts of the government testing capability and control measures such as the nationwide social-distancing and the statewide stay-at-home or shelter-in-place orders that result in an exponential decrease in the human social and movement activities. Quantitatively, for any State, the collective effects of these measures can be described by two parameters: the exponential decay rates of the activities associated with interstate and intrastate travel, denoted by λ_*inter*_ and λ_*intra*_, respectively, where a larger rate corresponds to more stringent control measures. The values of the two parameters characterizing the control measures can be estimated based on the available epidemic data [24]. Another key parameter is the fraction of undocumented infections, denoted as η, the role of which in the COVID-19 epidemic has been studied recently in the framework of the closed system, five-state model [24]. The value of η is determined by the testing and surveillance capability of the State. For clarity, we organize our results in terms of different combinations of λ_*inter*_, λ_*intra*_, and η, leading to distinct epidemic scenarios under government imposed control measures.

The human movement data for the populations into and out of a given State suggest that the interstate and intrastate activity decay rates are approximately equal (Methods): λ_*inter*_ ≈ λ_*intra*_ ≡ λ. For choosing reasonable values of λ, we use data from China to gain insights. In particular, the control measures imposed by the Chinese government were extremely stringent, where the human social activities are reduced to about 20% of the normal level after seven days of lockdown [24], giving rise to λ ≈ 0.24. In the US, the government measures are not as stringent as in China, so we set λ= 0.18, corresponding to a seven day reduction of about 30% in the social activities from the beginning of the implementation of the control measures. For comparison, we also simulate the hypothetical case of more strict control measures: λ = 0.24. To set the possible values of η for New York and other States in the US, we note that in China, due to the widespread government actions with extensive and sufficient testing and surveillance capabilities, most cases of infection were claimed to have been detected and reported, leading to an extremely low value of η: η = 0.012 as reported by the Chinese government (a recent work [26] reporting a substantial fraction of undocumented infections based on data from the early phase of the epidemic in China notwithstanding). In the US, due to lack of sufficient testing, the value of η can be much larger. While it is not possible to obtain an accurate estimate of the value of η for the US, it is not unreasonable that it can be at least 50%. We thus simulate two situations: η = 0.5 and 0.8. Taken together, for each of the ten States and the US as a whole, we carry out systematic simulations for four combinations of (η, λ): (0.5, 0.18), (0.5, 0.24), (0.8, 0.18), and (0.8, 0.24).

We focus on the time evolution of three quantities that are the key dynamical variables of our coupled five-state model (Methods): (i) the population of the individuals in the “hidden” state who have been infected but who are asymptomatic or show only mild symptoms, denoted as *H*(*t*), (ii) the population of infected individuals who show clear symptoms, denoted as *I*(*t*), and (iii) the population of confirmed individuals, denoted as *J*(*t*). In general, *H*(*t*) and *I*(*t*) exhibit a “humped” structure: with time they increase from zero, reach a maximum, and then decay to zero. The inflection point occurs when *I*(*t*) reaches maximum, and the epidemic is deemed over when *I*(*t*) has approached zero. The number of confirmed cases, *J*(*t*), is a non-decreasing function of time and reaches a constant value as *I*(*t*) approaches zero. The epidemic duration is measured by the date on which the H-state and I-state populations reach zero. The epidemic size can be characterized by the constant value of the final number of confirmed cases. To make quantitative predictions, it is necessary to have the values of the initial number of individuals in the hidden state and the infection rate, i.e., the probability for an individual in the susceptible state to switch to the *H* state, which can be estimated through an optimization procedure [24]. The values of the two parameters so obtained are denoted as *H*^*^ (0) and *β*^*^.

### Epidemic scenario for the State of Arizona

The starting date of COVID-19 epidemic in Arizona was March 5 (excluding a few sporadic cases before this date). The State population is about 7.28 millions. Figure 1(a) shows that, for η = 0.5 and λ = 0.18, the inflection point should occur on April 5 with the peak *I* value of about 2,600 and the epidemic will end around September 20 with 5,200 final confirmed cases. For the same value of η but λ = 0.24, the inflection point would occur three days earlier with the peak *I* value reduced by about 600, as shown in Fig. 1(b). In this case, the epidemic will be over three months earlier: on June 20, with 2,800 final confirmed cases - a reduction of about 46%. The results for η = 0.8 and λ = 0.18 are shown in Fig. 1(c): inflection point occurring on April 7, peak *I* value of about 2,800, ending date of epidemic around the middle of March 2021, and with about 8,800 final confirmed cases (one fifth of the actual infections). For η = 0.8 but λ = 0.24, the inflection point would occur around April 3 with the peak *I* value of 2,100, as shown in Fig. 1(d). Comparing with the case in Fig. 1(b) where tests are more extensive (λ = 0.5) but the restriction measures are more strict (η = 0.24), we see that, even when the test scale is significantly reduced (λ = 0.8), if more stringent control measures are imposed, the epidemic would only be slightly worse: delay of inflection by one day, increase in the peak *I* value by 100, one month longer in duration, and an approximate 17% increase in the number of final confirmed cases. Comparing with the real data of daily number of confirmed cases, we see that scenario (c) is the most likely current scenario for Arizona. Overall, the results for Arizona indicate the need to impose the strictest possible control measures, especially when extensive testing is not available.

**FIG. 1.**
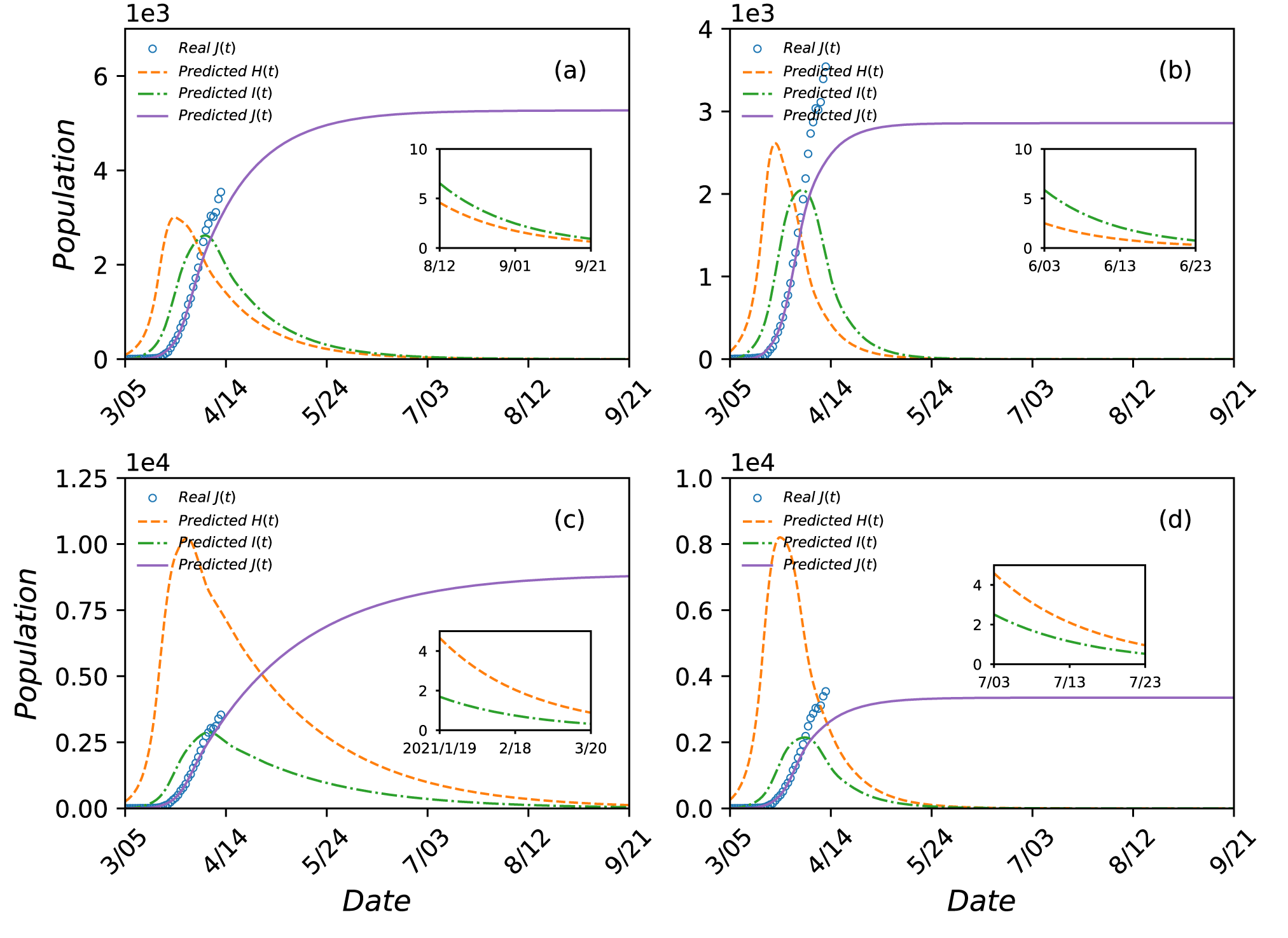
Predicted COVID-19 epidemic scenarios in Arizona as an open system. Shown are *H*(*t*) (dashed orange curve), *I*(*t*) (green dot-dashed curve), and *J*(*t*) (solid purple curve) from March 5 to September 21, 2020 predicted by the coupled dual system, five-state model. The parameter settings are (a) *η* = 0.5 and λ = 0.18, (b) *η* = 0.5 and λ = 0.24, (c) *η* = 0.8 and η = 0.18, and (d) *η* = 0.8 and λ = 0.24. For (a) and (b), optimization of the model equations gives *H*^*^(0)= 90 and *β*^*^ = 0.27. For (c) and (d), the corresponding values are *H*^*^(0)= 260 and *β*^*^ = 0.24. The inset in each panel shows the predicted *H*(*t*) and *I*(*t*) towards the end of the epidemic. The open blue circles are the actual data of *J*(*t*) available up to the time of writing. A comparison between the predicted and actual daily number of confirmed cases indicates (c) as the most likely scenario for Arizona at the present, where 80% of the cases remain unchecked and the government control measures are not as strict.

If Arizona was closed to traffic into and out of the State, simulation results (SN 2) reveal similar effects of the population movements on the epidemic quantities as those for the State of New York: small changes in the inflection occurrence time and peak *I* value but non-negligible impacts on the epidemic duration and the final infection size. For example, for the most likely current scenario (i.e., η = 0.8 and λ = 0.18), interstate travel will make the epidemic about half month longer and increase the final number of confirmed cases by about 1,200. This means that, even for Arizona where the epidemic is relatively mild, interstate travel poses a not-so-insignificant burden in State’s efforts to control the epidemic.

### Epidemic scenarios for the State of Washington

The State of Washington is unique because the first case of COVID-19 in the US occurred there and it was the epicenter in the early phase of the epidemic. However, the spreading subsided and stabilized quickly while outbreaks in other parts of the country occurred. Our model correctly predicts this rather unusual behavior, as follows.

In January and through most of February, there were a few cases in Washington State, a “non-epidemic” situation to which our model is not applicable. We use the daily number of confirmed cases from February 26 to March 29 to estimate the model parameters. The State population is approximately *N* = 7.6 × 10^6^. Figure 2 shows four distinct epidemic scenarios. In particular, Fig. 2(a) shows that, for η = 0.5 and λ = 0.18, the inflection point will occur on about April 3 with the peaked infected population of about 8,000. The epidemic will last through August 2 and the final number of confirmed cases will be about 13,000. For the same value of η (0.5), if the government measures are more stringent (λ = 0.24), as shown in Fig. 2(b), the occurrence of the inflection point will be two days earlier, the peak value of *I*(*t*) would be about 7,500, the epidemic would be approximately two months shorter (ending date around June 10), and about 11,000 cases will be finally confirmed - a 15% decrease as compared with the case of λ = 0.18 in Fig. 2(a). Now assume much reduced government testing and surveillance, resulting in η = 0.8. Figure 2(c) shows that, for λ = 0.18, the inflection point will occur on April 4 with the peak *I*(*t*) value of 9,000, the ending date of the epidemic will be about December 4, and the final size of the confirmed population will be 17,000 (so altogether 85,000 people would be infected). Now suppose a more restrictive set of measures: λ = 0.24. In this case, the inflection point will occur on April 2. Comparing with the case in Fig. 2(b), we see that the peak *I* value of 8,500 represents only a small increase, and the epidemic duration will be about one month longer (ending date on July 3) with 12,500 confirmed cases.

**FIG. 2.**
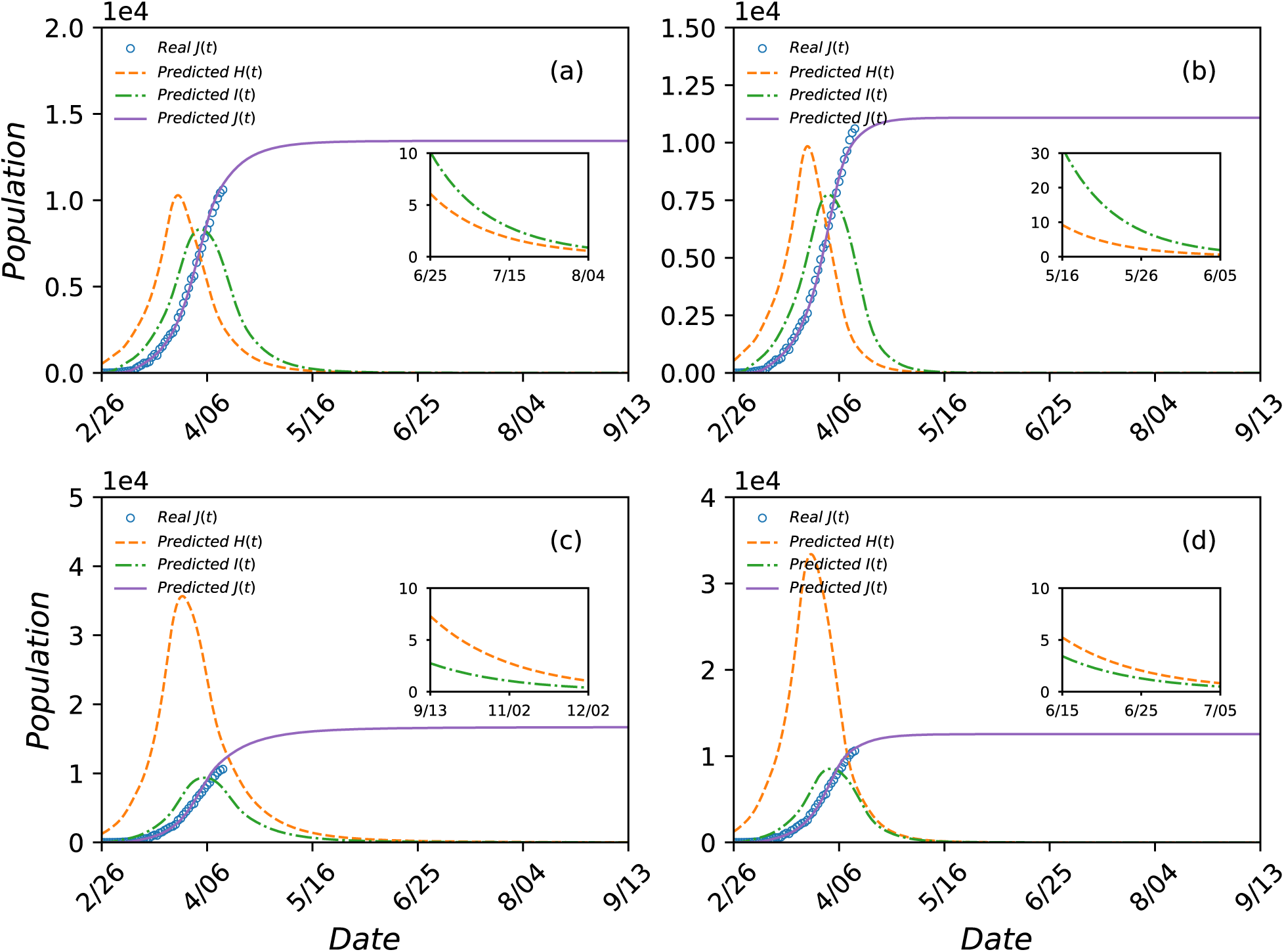
Predicted COVID-19 epidemic scenarios in Washington State as an open system. Legends are the same as those in Fig. 1. The time period covered is from February 26 to September 13, 2020. The parameter settings are (a) *η* = 0.5 and λ = 0.18, (b) *η* = 0.5 and λ = 0.24, (c) *η* = 0.8 and λ = 0.18, and (d) *η* = 0.8 and λ = 0.24. For (a) and (b), optimization of the model equations gives *H*^*^(0)= 530 and *β*^*^ = 0.18. For (c) and (d), the corresponding values are *H*^*^(0)= 1250 and *β*^*^ = 0.16. The optimally estimated values of the infection rate are markedly smaller than those of New York as indicated in the caption of Fig. 3. The inset in each panel shows the predicted behaviors of *H*(*t*) and *I*(*t*) towards the end of the epidemic. A comparison between the predicted and actual daily number of confirmed cases indicates (a) and (d) as the most likely scenarios for Washington at the present.

Comparing the projected *J*(*t*) with the current data, we find that Figs. 2(a) and 2(d) represent the most likely scenarios for State of Washington at the present. This is intriguing because the two cases differ markedly in terms of the State testing capability and strength of control measures. Another result is that the epidemic scale of this early epicenter is projected to be at least one order of magnitude smaller than that of the current epicenter New York. A plausible explanation for these phenomena lies in the relatively low value of the infection rate obtained through data based optimization (Methods): β^*^ = 0.18 for Figs. 2(a) and 2(b), and β^*^ = 0.16 for Figs. 2(c) and 2(d), which should be compared with the respective values of about 0.25 for other States. The “abnormally” low values of β, which may result from relatively low level of human activities in the State of Washington, make the impacts of testing and control measures less significant in comparison with other States.

### Epidemic scenarios for the State of New York

At the time of writing, New York State is the epicenter of COVID-19 in the US with the largest number of confirmed cases and most deaths among all 50 States. The starting date of simulating the epidemic in the State is March 2 and the population of the State is approximately 19 millions. Figure 3 presents the predicted outcomes of four scenarios. In Fig. 3(a), the fraction of undocumented infection is 50% (η = 0.5) and the control measures imposed by the State government are relatively less stringent (λ = 0.18). In this case, the inflection point will occur around April 8 when the infected population *I*(*t*) is peaked at the value of approximately 300,000. Because the mean time delay from I state to J state is set as seven days, the population of newly confirmed patients will be peaked around April 15, the epidemic will be over on about November 21, and the final number of confirmed cases would be about 550,000. In Fig. 3(b), η is still 50% but more stringent control measures are assumed: λ = 0.24. In this case, the inflection point will occur on about April 5 with the peaked value of *I*(*t*) about 240,000. Under the stringent measures, the epidemic will be over around August 1 with the final number of confirmed cases about 330,000. Figure 3(c) shows that, for η = 0.8 and λ = 0.18, the inflection point will occur around April 9 with the peaked infected population 350,000. The final number of confirmed cases would be 750,000 and the epidemic would last into 2021 to end around May 30. Because only 20% of the infections are documented, the actual infected population would be about 3.75 millions. Figure 3(d) shows that, for η = 0.8 and λ = 0.24, the inflection point will occur around April 6 with the peaked infected population 270,000. In this case, the epidemic will last until September 6 with the total number of confirmed cases about 400,000. Comparing Fig. 3(c) with Fig. 3(a), we see that, if the control measures are not as stringent (λ = 0.18), insufficient tests and surveillance leading to an increase in the value of h from 50% to 80% will have devastating consequences: the epidemic duration will be significantly longer (six more months) and the number of confirmed cases will be 36% higher. However, if the government measures are stringent (λ = 0.24), as can be seen by comparing Figs. 3(d) with Fig. 3(b), the same increase in the value of η would result in only one extra day in the arrival of the inflection point, a month delay in the duration, and about 20% increase in the number of confirmed cases. These results indicate that, relative to testing and surveillance, imposing more stringent control measures would be more effective at containing the disease spreading.

**FIG. 3.**
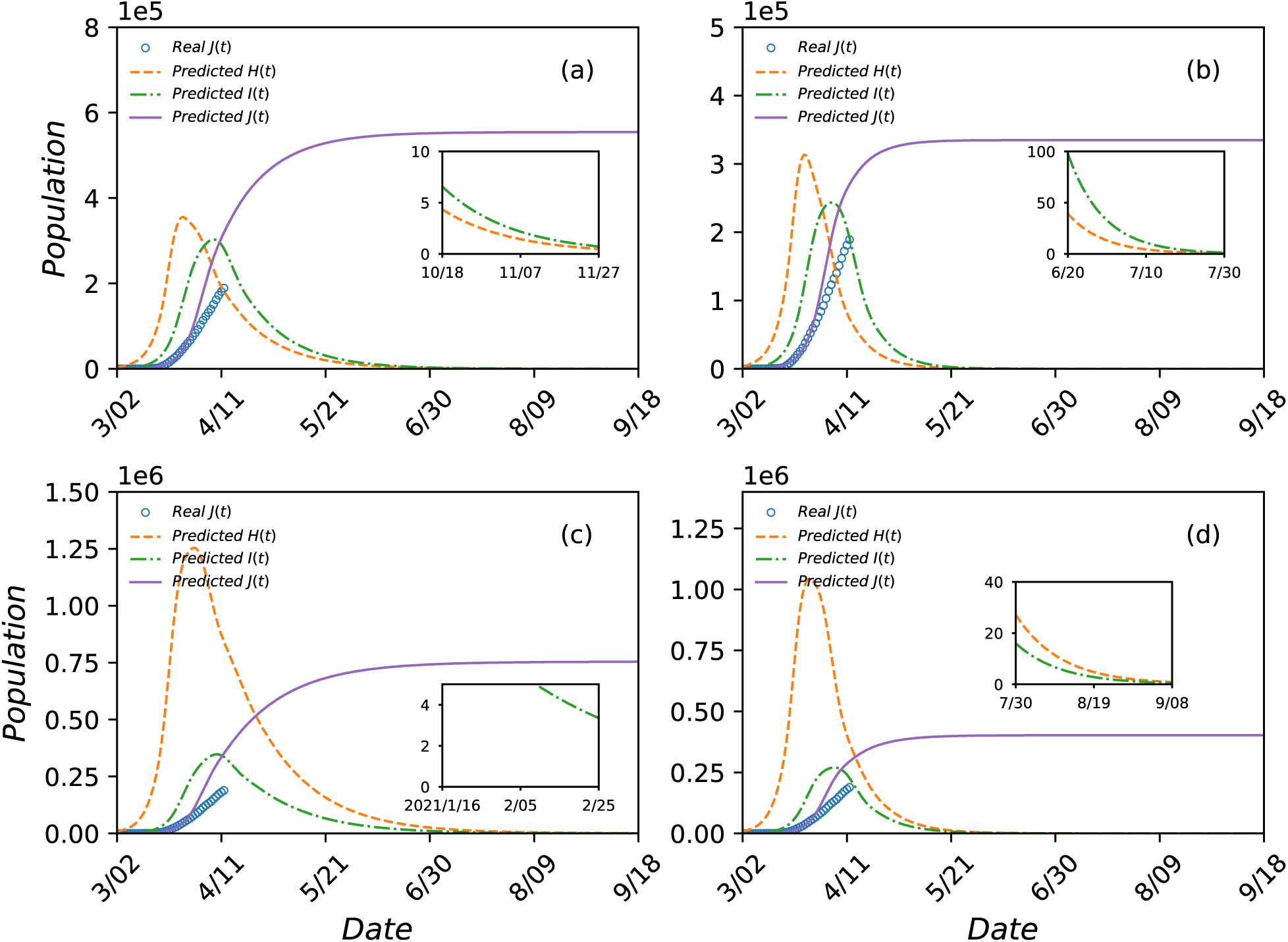
Prediction of COVID-19 epidemic scenarios in New York State. Legends are the same as those in Fig. 1. The parameter values are: (a) η = 0.5 and λ = 0.18, (b) η = 0.5 and λ = 0.24, (c) η = 0.8 and λ = 0.18, and (d) η = 0.8 and λ = 0.24. For (a) and (b), the optimal estimates of the number of initial hidden population and the inflection rate are *H*^*^(0)= 3400 and *β*^*^ = 0.27. For (c) and (d), these values are *H*^*^(0)= 8600 and *β*^*^ = 0.25. The inset in each panel shows the diminishing trend of *H*(*t*) and *I*(*t*) during the last stage of the epidemic. A comparison of the model generated *J*(*t*) with the real data indicates that none of the scenarios fits with the current trend in the State of New York. See text and SN 3 for an explanation.

Checking against the available data to date, we find that none of the four scenarios in Fig. 3 matches the real COVID-19 trend in the State of New York. This is due to a highly localized or “singular” behavior: most infections in the State have occurred in New York City. Indeed, if we treat New York City as an independent “State” system, a good match between some predicted scenario(s) and the empirical data arises. Detailed results are presented in SN 3.

Because of the severity of the epidemic in New York, it is of interest to assess the impact of control measures in a systematic manner. Because these measures are imposed by the State government, we consider the State of New York and calculate the epidemic duration and the final sizes of the three key populations versus λ. Figure 4(a) shows, for η = 0.5, the epidemic ending date versus λ. It can be seen that strengthening the measures can lead to an exponential shortening in the duration of the epidemic. The corresponding results for η = 0.8 are shown in Fig. 4(b). Figures 4(c) and 4(d) show, for η = 0.5 and η = 0.8, respectively, the peak *H* and *I* populations as well as the final size *J* of the confirmed cases versus λ, where more stringent government measures can lead to an exponential decrease in the final size of the confirmed population. In terms of specific numbers, a loose set of control measures, e.g., λ= 0.16, will result in 800,000 (η = 0.5) or 1.1 million (h = 0.8) confirmed cases for the State of New York. However, a stringent set of measures, say λ = 0.28, would reduce the corresponding number to about 300,000 for both η = 0.5 and η = 0.8. That is, if the government measures are sufficiently strict, the size of the tested population becomes an insignificant factor determining the epidemic trend.

**FIG. 4.**
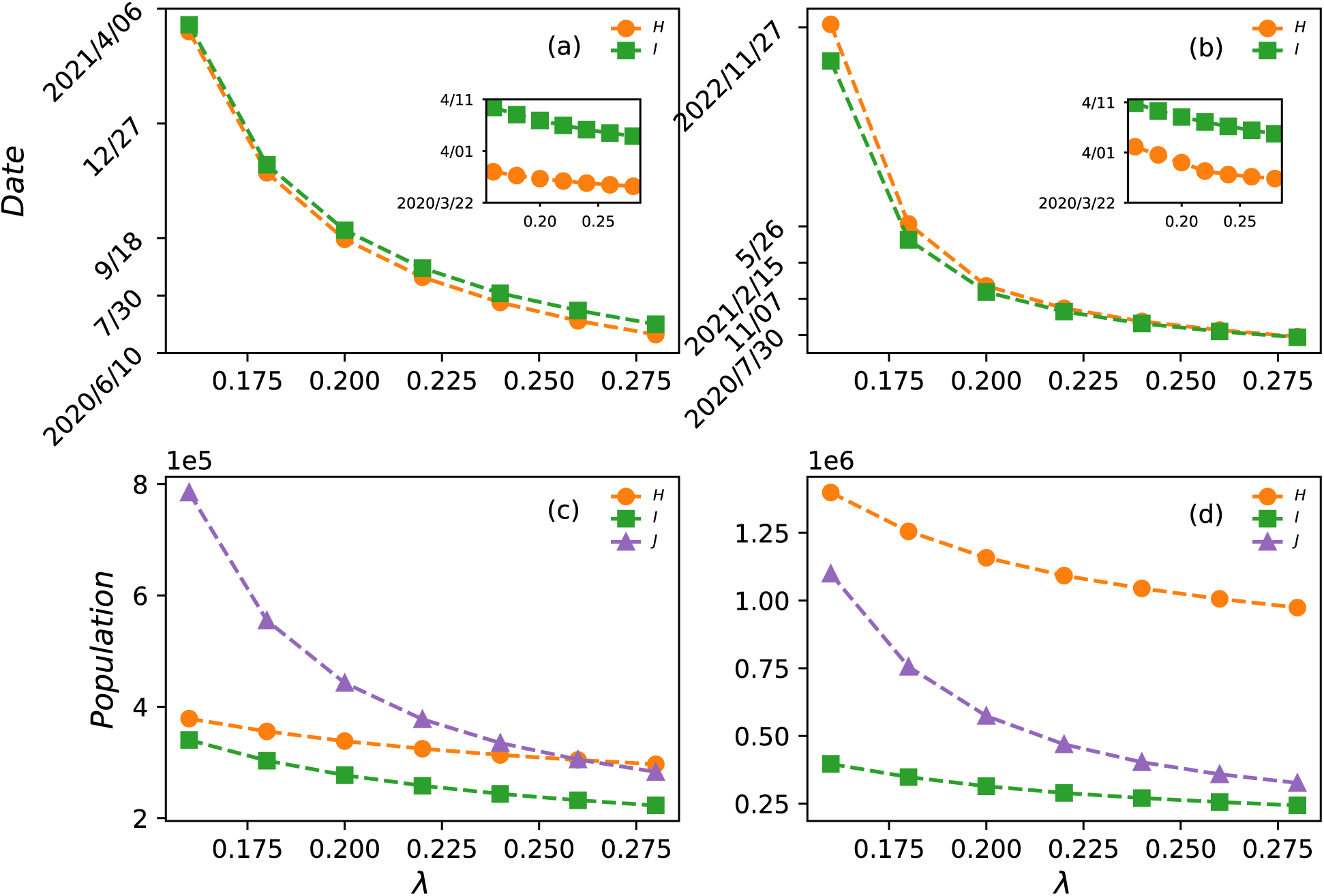
Effect of varying government control measures on epidemic suppression in New York State. (a,b) For η = 0.5 and η = 0.8, respectively, the ending date versus λ. (c,d) For h = 0.5 and η = 0.8, respectively, the peak populations of *H* and *I* as well as the final population *J* of confirmed cases versus λ.

How do the daily population movements into and out of the State affect the epidemic trend? To address this question, we simulate the closed five-state model for the State of New York and compare the results (SN 4) with those in Fig. 3. We find that the population movements have relatively small effects on the occurrence time of the inflection point and the peak value of the *I*-state population, but can have a dramatic effect on the epidemic duration and the final size. For example, for η = 0.8 and λ = 0.18, the population movements can make the duration four months longer but can reduce the final size of the confirmed population by about 15,000. This means that, without a complete lockdown of the entire State of New York, the travelers out of the State will spread the virus to the rest of the country, leading to a prolonged epidemic duration for the US and the State itself. This conclusion is also supported by simulation results of the closed model but for the entire country (SN 5), which demonstrate a similar epidemic trend as that of New York for every parameter setting tested. The implication is that, while COVID-19 has already begun to spread in other States throughout the country, a complete lockdown of the epicenter would still be beneficial or even necessary. Merely practicing social distancing and imposing stay-at-home order are not sufficient!

### Effects of interstate and intrastate travel restrictions

The results obtained so far are under the assumption that interstate and intrastate travel restrictions generate the same rate of reduction in the social activities: λ_*inter*_ = λ_*intra*_. In reality the two rates can be different. To systematically assess the effect of this difference, we fix one rate and calculate the epidemic duration and size as a function of the other rate for the States of New York and Arizona. Figures 5(a) and 5(b) show, for λ_*intra*_ = 0.18 and λ_*inter*_ = 0.18, respectively, the ending date of the epidemic in New York versus λ_*inter*_ and λ_*intra*_. The corresponding results for Arizona are shown in Figs. 5(c) and 5(d). The inset in each panel shows the date of the occurrence of the inflection point. From Figs. 5(a) and 5(c), we see that tighter restrictions on the interstate traffic leading to an increase in the value of λ_*inter*_ affect little the occurrence of the inflection point but have some effect on the epidemic duration. Because of the ongoing large scale outbreak in New York, imposing heavier restriction on interstate traffic only has an incremental effect on reducing the duration, as shown in Fig. 5(a). However, for Arizona, because the outbreak is not severe, a stronger restriction on interstate traffic is more effective at shortening the epidemic, as shown in

**FIG. 5.**
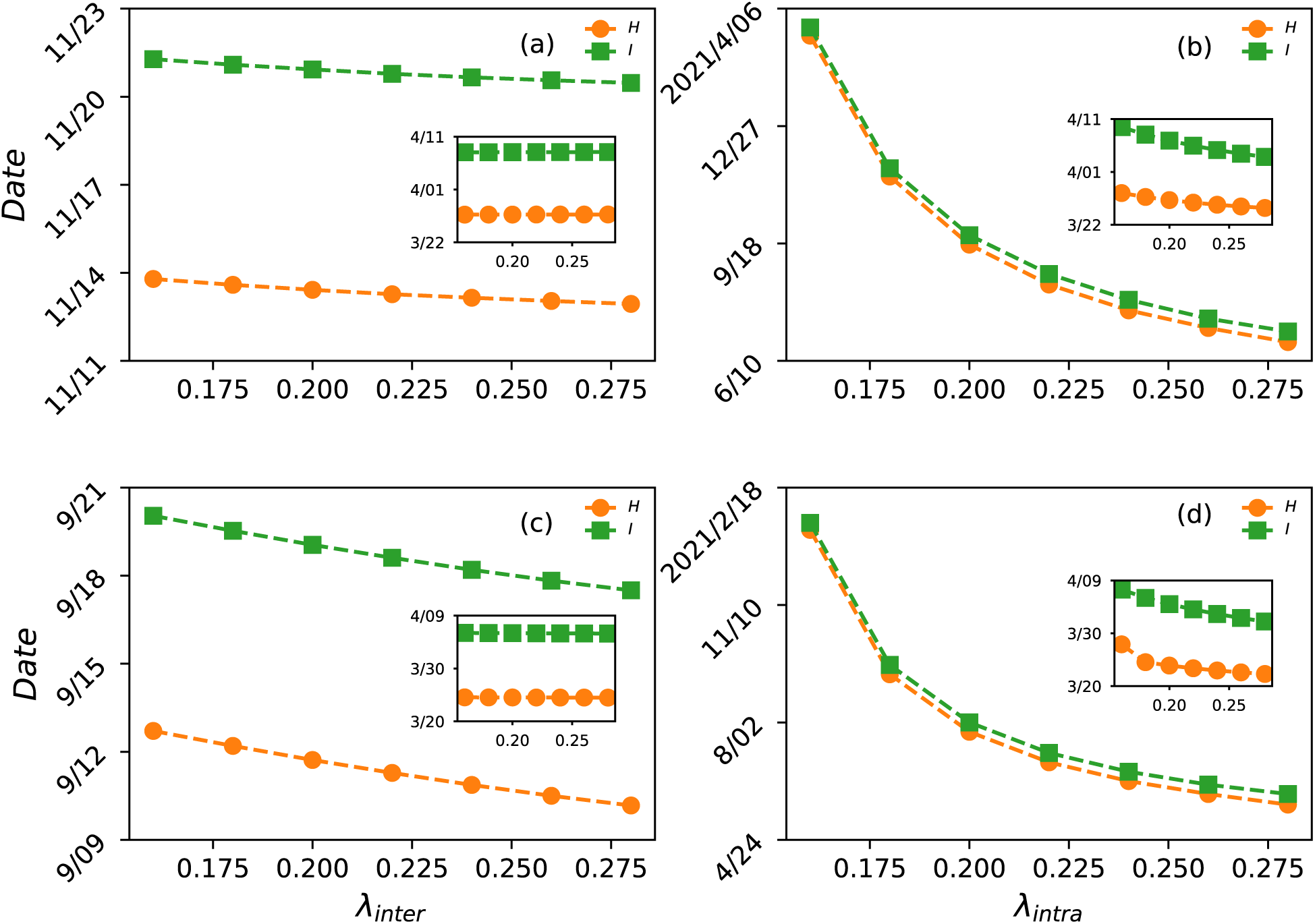
Impact of interstate and intrastate traffic restriction on epidemic duration in New York and Arizona. (a) For New York, the ending dates of H (orange) and I (green) versus the interstate traffic reduction rate λ_*inter*_ for λ_*intra*_ = 0.18. (b) For New York, the ending dates versus the intrastate traffic reduction rate λ_*inra*_ for λ_*inter*_ = 0.18. In (a,b), the fraction of undocumented inflections is η = 0.5 and the optimized model parameter values are *H*^*^(0)= 3400 and *β*^*^ = 0.27. (c,d) The corresponding results for Arizona for η = 0.5, *H*^*^(0)= 90, and *β*^*^ = 0.27. For both States, the effects of interstate traffic restriction are relatively small (a,c), but the intrastate traffic restriction leads to an exponential shortening of the epidemic duration. Inset: date of occurrence of inflection point in terms of the peak values of *H* (orange) and *I* (green).

Fig. 5(b). For both New York and Arizona, tightening the restriction on intrastate traffic can have an exponential effect on shortening the duration, as shown in Figs. 5(b) and 5(d), respectively, although the effect on advancing the occurrence of the inflection point is small (insets). Similar results have been obtained on the effects of the interstate or intrastate traffic restrictions on the epidemic size for New York and Arizona (SN 6). These results justify and demonstrate the benefits for the State government to issue strict stay-at-home or shelter-in-place orders to limit intrastate activities.

### Epidemic scenarios for seven other States

Predicted scenarios for seven other States: New Jersey, California, Michigan, Florida, Illinois, Massachusetts, and Louisiana are presented in SNs 7-13, respectively. The results for the ten States and the US are summarized in Supplementary Table 1.

## DISCUSSION

The number of confirmed cases in the United States has exceeded a quarter of global cases and the epidemic is evolving rapidly in some States. Unlike countries such as China where the orders of the central government are uniformly and strictly followed in the entire country, there is vast heterogeneity among the States in the US in terms of population movements and the government restriction orders, leading to drastically varying epidemic dynamics in different States. As a result, prediction of the COVID-19 epidemics in the US needs to be done in a State-by-State manner. A complication is that, because of the daily interstate population movements, each individual State is effectively an open system, rendering inapplicable the recently developed non-Markovian model for a closed system [24]. We have developed a generalized, non-Markovian epidemic model for COVID-19 in an open setting, taking into account population movements into and out of the system.

The main goal is to predict, for each of the ten States selected for this study (the nine States with the most severe COVID-19 epidemics in the early stage, plus the State of Arizona), the most likely epidemic scenario(s) based on the currently available data. In particular, for any given State, we have used the daily number of confirmed cases up to March 29 to estimate the two key model parameters: the initial population of hidden state and the infection rate, enabling the model to generate possible epidemic trajectories into the future. Comparing the trajectories with the available data up to the time of writing (April 12) allows us to identify the most likely epidemic scenario(s) for the State. For all ten States except the State of New York, at least one such epidemic scenario can be identified. The difficulty with New York State has been resolved by noting the existence of a singular and highly localized spot: New York City, into which vast majority of the cases in the State fall, generating an exceptionally high degree of heterogeneity in the State and rendering inaccurate model prediction of the entire State. The situation is similar to attempting to predict the possible epidemic scenarios for the entire country of USA, which is not feasible due to the highly heterogeneous pattern of outbreaks. Indeed, simulation of New York City alone as an independent system has generated two most likely scenarios that match with the data. The difficulty with New York State and our successful resolution have thus provided further justification for the State-by-State prediction approach. All these results have not only validated our model, but also demonstrated its predictive power. The most likely epidemic scenario(s) for any given State is (are) indicative of whether testing services and control measures are vigorous enough to fully contain the spreading of COVID-19, providing guidance for improvement.

Another goal is to evaluate the effects of government imposed control measures on the epidemic trends through various scenarios. We have reported results from model predictions of the ten States that differ in their testing and surveillance capabilities. The measures imposed by the States to control COVID-19 also vary, which can be characterized by the exponential rates of reduction in the interstate and intrastate human activities. Our finding is that, with insufficient testing capability leading to a larger fraction of undocumented cases (e.g., 80% versus 50%) and with strict control measures, the occurrence of the inflection point will be delayed for a few days, the peak infected and documented population will increase by 20%, the epidemic duration will be prolonged for as long as half year, and the final infected population can increase by 70%. However, relaxing the control measures can have more devastating consequences. For example, a loose restriction measure (e.g., social-distancing only without stay-at-home order) can lead to a delay in the occurrence of the inflection point by ten days, a 40% increase in the peak infection, epidemic duration of over one year, and a more than five-fold increase in the size of the final infections (documented or undocumented). Simulation results incorporating uncertainties in the prediction still support the above conclusions (SN 13). We also find that imposing intrastate movement restriction can suppress exponentially the epidemic in both its duration and size, regardless of the current epidemic trend of the State. A comparison among the predicted scenarios for different States reveals a difference in the occurrence of the inflection point of about ten days but a stretch in the epidemic duration for as long as one year. Especially, simulations treating the State of New York as an open or a closed system reveal that, if the system is open, the final confirmed cases would be reduced by 15,000 but the duration can be longer by four months. This is because, a fraction of the reduced infected population in New York would diffuse into other States, making the epidemic of the whole country longer. Interstate travel thus poses a significant threat to States such as Arizona, where the epidemic is much less severe. This suggests the necessity of a complete lockdown of New York, the current epicenter, to significantly shorten the epidemic in the US as a whole. Our findings suggest that the duration of such strict lockdown should be no less than one year.

## METHODS

### Non-Markovian, five-state model for COVID-19 in any individual State as an open system

To model the COVID-19 epidemic in an open system, three unique features must be considered: (1) undocumented population with no or mild symptoms, (2) non-Markovian state transitions, and (3) time dependence due to human movements in and out of the system. Models developed in the past few months [2–9, 27] provide insights but do not take into account the three features. If the system is closed, it is only necessary to consider the first two features, resulting in a five-state model [24]. Here we generalize the closed-system model to include time dependence.

Figure 6 illustrates the generalized model. An individual can be in one of the five states at each time step: susceptible (S), hidden (H), infected (I), confirmed and isolated (J), and removed (R). The states S, I, and R have the same meanings as in the classical SIR model for infectious disease, but states H and J are unique for COVID-19. In particular, an individual in H has had the virus and is infectious but is asymptomatic or only mildly symptomatic, in contrast to the I state in which individuals show symptoms. The J state contains individuals who are confirmed with COVID-19. Note that, individuals in the I or J state are quarantined or hospitalized, so the probability for them to infect others can be neglected. Individuals in the R state, by definition, are not infectious. Thus, only the H individuals are capable of infecting others [24].

**FIG. 6.**
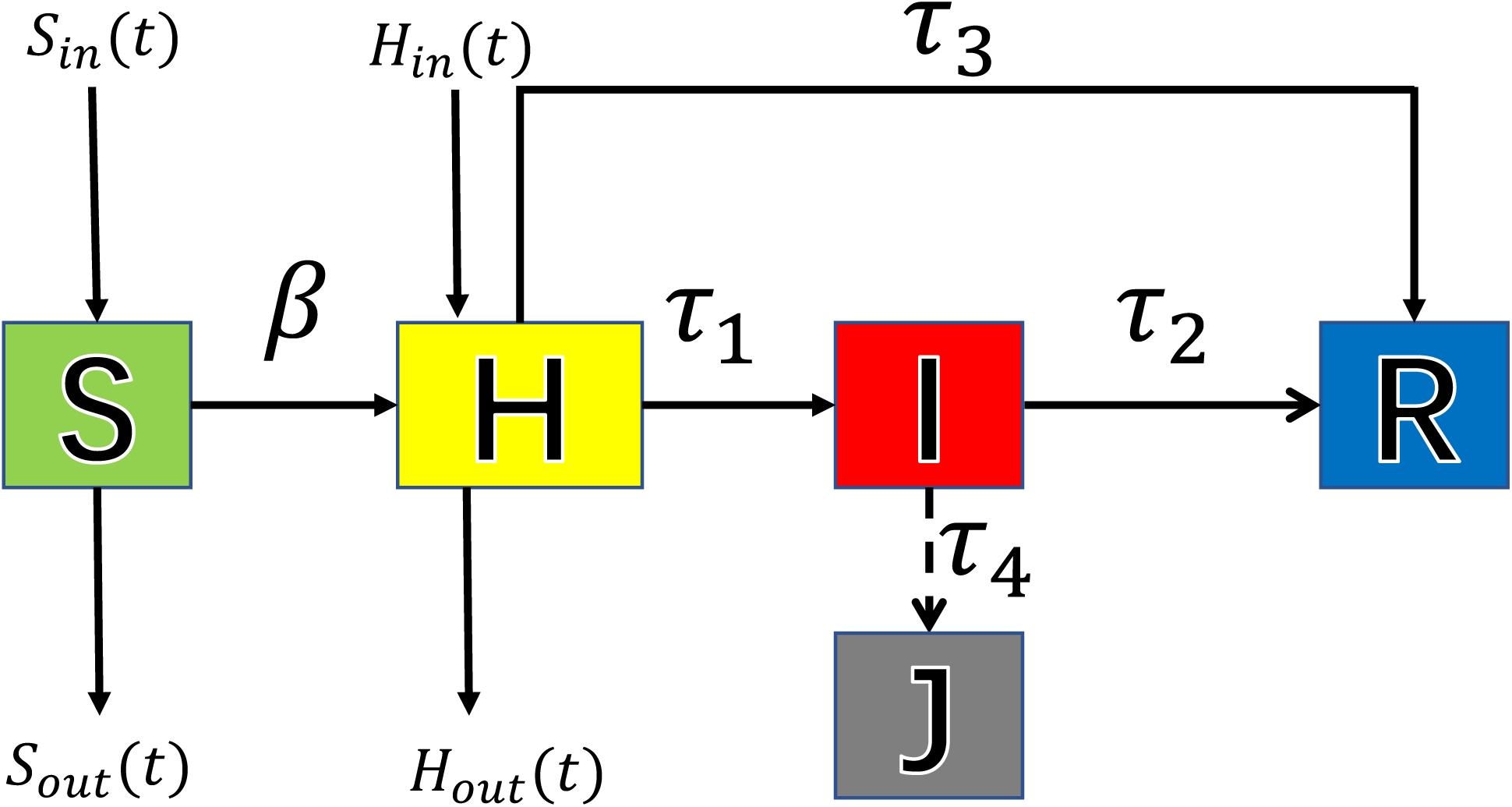
Schematic illustration of generalized COVID-19 model for an open system. The five states are: S (susceptible), H (hidden), I (infected), J (infected and confirmed), and R (removed: cured or died). Parameter *β* is the infection rate, and *τ*_1_, *τ*_2_, *τ*_3_, and *τ*_4_ represent the four relevant time delays between different state pairs due to the non-Markovian nature of the epidemic. The four quantities *S*_*in*_(*t*), *S*_*out*_ (*t*), *H*_*in*_(*t*), and *H*_*out*_ (*t*) represent the populations moving into and out of the S and H states of the open system.

The spreading dynamical process can be described, as follows. Individuals in the S state are infected by H individuals at the rate b and switch to the H state. A fraction h of H-state individuals recover spontaneously or die, a process that requires *τ*_3_ days. The parameter h thus represents the fraction of undocumented infections, and its value is determined by the testing capability of the country or State. The remaining (1 − η) fraction of H individuals go through a transition to the I state after an average incubation period of *τ*_1_ days - a typical non-Markovian process. With medical treatment, individuals in the I state recover or die after *τ*_2_ days. Finally, a time delay exists for the transition from I to J: on average the I individuals will need *τ*_4_ days to be confirmed.

We treat each State in the US as an open system, regarding the influences from all the other States as perturbations, mathematically represented by the populations moving into and out of the S and H states, denoted as *S*_*in*_(*t*), *S*_*out*_ (*t*), *H*_*in*_(*t*), and *H*_*out*_ (*t*), respectively, as shown in Fig. 6. These functions are determined by the travel intensity as a function of time. Two types of travel need to be distinguished: interstate and intrastate, with the corresponding intensity functions *l*_*inter*_(*t*) and *l*_*intra*_(*t*). Due to government imposed travel restrictions, these functions decay exponentially from an initial value to a final smaller constant value. For instance, a recent estimate [15] gives that, for several major US cities, the travel restrictions would reduce the outbound human movements by 50%. In general, we have

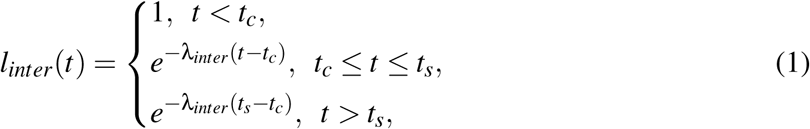

and

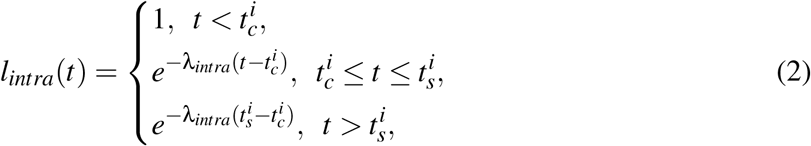

where *t*_*c*_ and 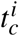 are, respectively, the starting dates of interstate and intrastate travel restrictions and the exponential decay in the movement activities occurs between *t*_*c*_ and *t*_*s*_ or between 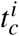 and 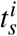. The starting dates differ from State to State. For the ten States studied, the dates 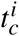 are listed in Supplementary Table 2. The values of *t*_*s*_ and 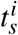 are set as *t*_*c*_ + 7 and 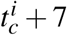, respectively. Our generalized five-state model for COVID-19 epidemic for any given target State in the US (an open system) can be described by the following set of delayed integro-differential equations:

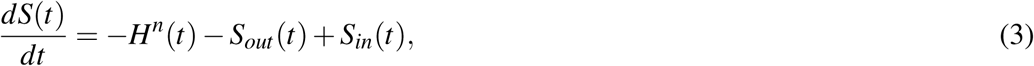

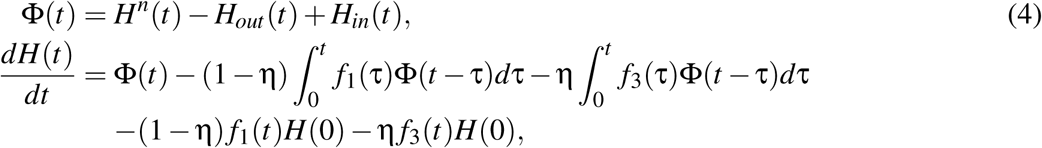

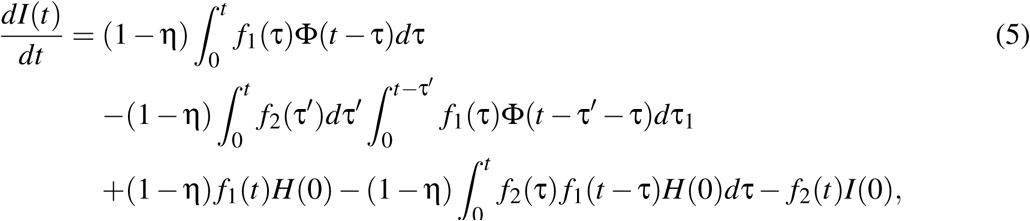

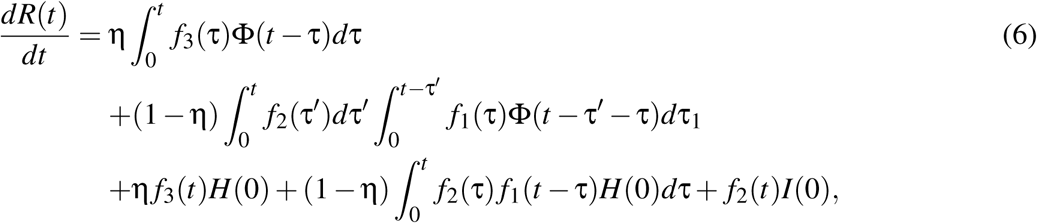

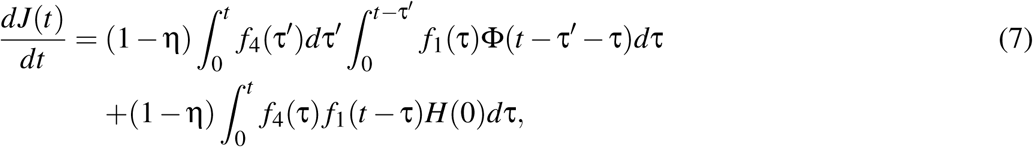

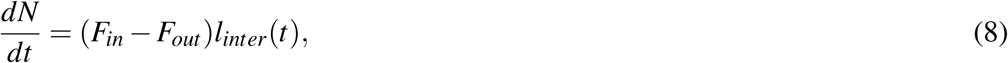

where the quantity *H*^*n*^(*t*) in Eqs. (3) and (4) is the rate of increase in the H-state population: *H*^*n*^(*t*)= β*S*(*t*)*l*_*intra*_*H*(*t*)*/N*(*t*) with *l*_*intra*_(*t*)*H*(*t*) representing the active H-state population that has not been isolated, *f*_1_(*τ*), *f*_2_(*τ*), *f*_3_(*τ*), and *f*_4_(*τ*) are the normal probability distribution functions [24] of the delay time *τ*_1_, *τ*_2_, *τ*_3_, and *τ*_4_, respectively, and *N*(*t*) is the population of the State as a function of time. The quantity Φ (*t*) is the increment of the H-state population. A term-by-term explanation of the integro-differential equations can be found in Ref. [24]. The input and output functions given by

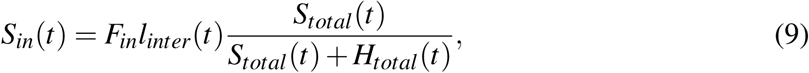

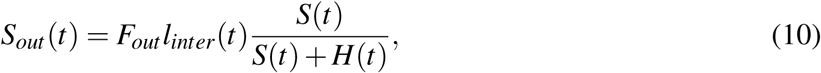

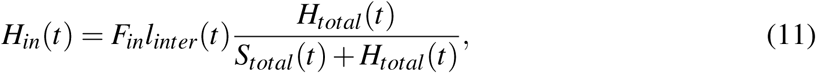

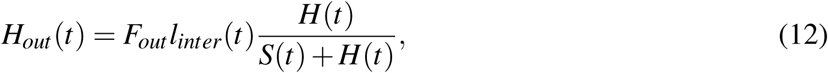

where the quantities *S*_*total*_(*t*) and *H*_*total*_(*t*) are the total S and H-state populations of the US excluding the target State, *F*_*in*_(*t*) and *F*_*out*_ (*t*) are the fluxes into and out of the State, which can be extrapolated from empirical data. To numerically solve the whole set of equations, the values of the initial H-state population, *H*(0), and of the infection rate β are needed, which can be estimated through a mathematical optimization procedure [24].

### Determination of in and out population fluxes

To determine the daily fluxes *F*_*in*_(*t*) and *F*_*out*_ (*t*), we use the commuting data from the US Census Bureau (https://www.census.gov/topics/employment/commuting.html), which were obtained from sampling the home and work addresses of the working population in the five-year period (2011-2015). Our estimation method is described, as follows. Assume that the commuting population is distributed uniformly among the States. On average, each working individual commutes 0.11 time per day. Multiplying this number by the population of the State gives the daily average number of people who commute. Denoting this number by *Q* and letting the commuting populations from the Census Bureau’s data base be *P*_*in/out*_, we obtain the ratio *C*_*in/out*_ = *Q/P*_*in/out*_. Let *D*_*in/out*_ be populations in and out of the State from the data base. The daily fluxes can be obtained as *F*_*in/out*_ = *C*_*in/out*_ · *D*_*in/out*_. For the ten States studied, the flux values are listed in Supplementary Table 3.

## Data Availability

All relevant data are available from the authors upon request.

## DATA AND CODE AVAILABILITY

All relevant data are available from the authors upon request. All relevant computer codes are available from the authors upon request.

## ACKNOWLEDGMENTS

We thank Prof. Xiaoyong Yan for suggestion on modeling population fluxes. This work was supported by the National Natural Science Foundation of China (Grant Nos. 11975099, 11575041, 11675056 and 11835003), the Natural Science Foundation of Shanghai (Grant No. 18ZR1412200), and the Science and Technology Commission of Shanghai Municipality (Grant No. 14DZ2260800). YCL would like to acknowledge support from the Vannevar Bush Faculty Fellowship program sponsored by the Basic Research Office of the Assistant Secretary of Defense for Research and Engineering and funded by the Office of Naval Research through Grant No. N00014-16-1-2828.

## AUTHOR CONTRIBUTIONS

M.T., Z.L., and Y.-C.L. designed research; Z.-M.Z., Y.-S.L., J.K., and Y.-L.L. performed research; L.Z., L.-L.H, Z.-H.L., Y.-Q.Z., D.-Y.W., and D.X. contributed analytic tools; Z.-M.Z., Y.-S.L., L.Z., M.T., D.X., and Y.-C.L. analyzed data; Y.-L.L., J.K., M.T., Z.L., and Y.-C.L. wrote the paper.

## Supplementary Information for

### Supplementary Note 1: Recent efforts in COVID-19 modeling and prediction

There have been focused recent efforts in predicting the epidemic trends of COVID-19 and the effects of government control measures through mathematical modeling [1–23]. For example, two control strategies, one focusing on mitigation and another on suppression, were studied and compared [11] with the conclusion that, in the United Kingdom, even some best and optimal mitigation measures can lead to a two-thirds reduction in the demand for medical diagnosis and treatment and a 50% decrease in the number of deaths, the epidemic would still result in hundred thousands deaths and overwhelm the intensive care system. The suggestion is that, even for countries whose health care systems are still functional at the present, the strategy of suppression should be given a higher priority. Another work studied the epidemic risks and trends in different cities in Spain [13] through traffic data-driven, stochastic simulations of virus propagation and diffusion in spatial regions and inference of the starting time of the epidemic. The results suggest that finding and isolating the infected individuals at the earliest possible time as well as taking intervention measures to reduce the infection rate are effective strategies. Simulations of traditional mathematical models in epidemiology suggest the benefits of local prevention measures such as forbidding mass social gatherings, pinning down the exposure time as an important factor in disease spreading [14]. For example, if the basic reproduction number is two and the infection period is 14 days, if an infected individual stays at a gathering for more than nine hours, the likelihood of infecting others is high. If the exposure time exceeds 18 hours, the participants in the gathering should take strict measures to protect themselves. The effects of travel restrictions on COVID-19 spreading were studied [15], with the finding that the lockdown of Wuhan city in China provided a golden time between three and five days for other regions of China to get prepared and, by the middle of February, might have suppressed the scale of global pandemic by as much as 80%. It was suggested that travel restriction in combination with individual prevention measures such as wearing facial masks leading to a reduction in the infection rate by more than 50% can suppress the epidemic. The exponential growth rate in different regions of China in the early stage of COVID-19 epidemic was estimated [16] through a significant level analysis, revealing an approximately uniform value for China: 2.1 *±* 0.3 with the finding that effective control strategies leading to dramatic changes in the collective behaviors of the vulnerable population have a direct impact on the growth rate. A network model based on dynamic compartments and the SEIR (susceptible-exposed-infected-recovered) spreading dynamics was articulated [18] to analyze the effects of travel restriction, business closure, and social distancing on the epidemic, with the finding that school closure would shield the youth population from the disease, bar closures would be beneficial to young people and adults alike, and canceling religious services would protect the adults and the elders.

Models for the COVID-19 epidemic in the United States have also been developed. A metapopulation model was proposed to simulate the SARS-CoV2 spread and the effects of social distancing and travel restriction on suppressing the epidemic in the continental US [20] with the prediction of large scale outbreak in the country within 180 days after March 13. A network-driven epidemic dynamic model was constructed to simulate the COVID-19 progression timeline and the effectiveness of interventions across the US [21] with the prediction that, without control measures and travel restriction, about 7% of the population of the country would be infected when the epidemic peaks at the beginning of June. If the infection rate of COVID-19 is weakened by 25%, the peak would be delayed for about 34 days and its height would be reduced by 39%. Simulations of a model adopted from influenza management suggested [11] the necessity to maintain the control measures until a vaccine becomes available in about 18 months, but socially and economically this would represent a great challenge. Intermittent implementation of social distancing according to epidemic development can allow relaxation of control measures in relatively short time windows: in case a second wave of epidemic begins to emerge, social distancing and family isolation of suspected infections can be put in place again with an increasing intensity. The growth rate and reproductive number of COVID-19 were estimated [22] between March 14 and 19 for different cities in the US, revealing a power law relationship between these quantities and the city population. The implication is that the spreading speed in larger cities will be higher and, without control and prevention measures, a larger fraction of the population would be infected in larger cities. A dynamic model for COVID-19 was proposed [23] for comparing containment strategies in a pandemic scenario that the government can choose: one is a persistent control measure and another is based on adaptive implementation of suppression strategies with intensity depending on time, where the latter can shorten the epidemic duration to a greater extent as compared with the former. In fact, more lives can be saved if strict control measures are implemented earlier.

### Supplementary Note 2: Epidemic scenarios in Arizona as a closed system

We simulate four distinct epidemic scenarios for the State of Arizona treated as a closed system with different combinations of testing capability and strength of control measures. The results are listed in the table below.

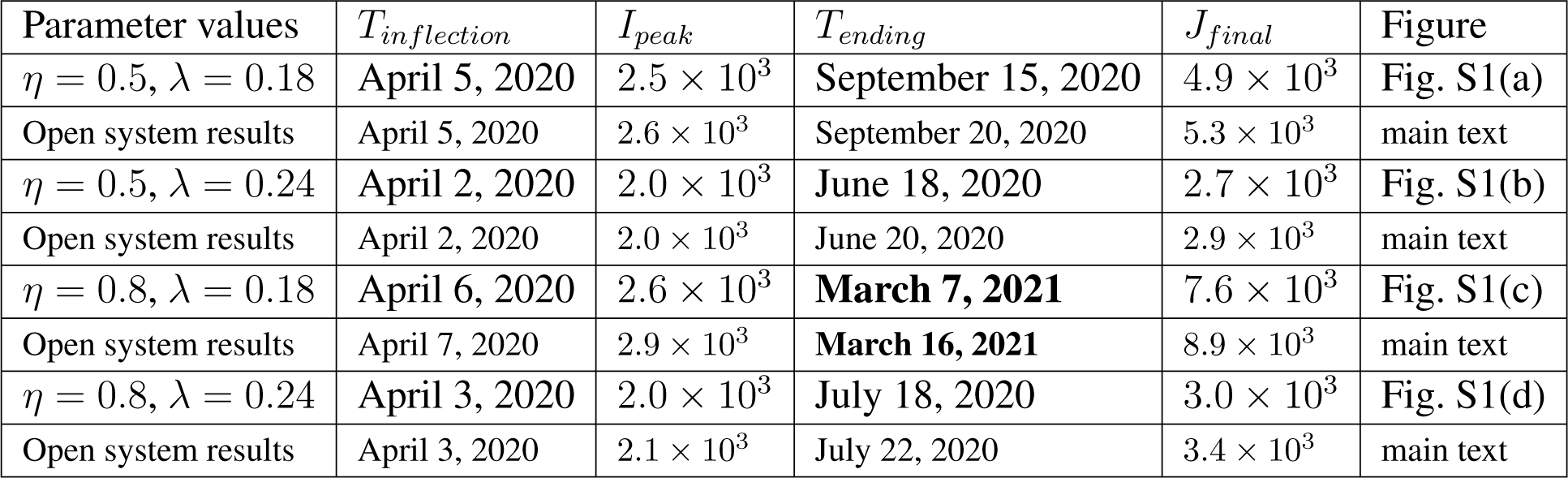

| Parameter values | $T_{inflection}$ | $I_{peak}$ | $T_{ending}$ | $J_{final}$ | Figure |
| --- | --- | --- | --- | --- | --- |
| $\eta = 0.5, \lambda = 0.18$ | April 5, 2020 | $2.5 \times 10^3$ | September 15, 2020 | $4.9 \times 10^3$ | Fig. S1(a) |
| Open system results | April 5, 2020 | $2.6 \times 10^3$ | September 20, 2020 | $5.3 \times 10^3$ | main text |
| $\eta = 0.5, \lambda = 0.24$ | April 2, 2020 | $2.0 \times 10^3$ | June 18, 2020 | $2.7 \times 10^3$ | Fig. S1(b) |
| Open system results | April 2, 2020 | $2.0 \times 10^3$ | June 20, 2020 | $2.9 \times 10^3$ | main text |
| $\eta = 0.8, \lambda = 0.18$ | April 6, 2020 | $2.6 \times 10^3$ | **March 7, 2021** | $7.6 \times 10^3$ | Fig. S1(c) |
| Open system results | April 7, 2020 | $2.9 \times 10^3$ | **March 16, 2021** | $8.9 \times 10^3$ | main text |
| $\eta = 0.8, \lambda = 0.24$ | April 3, 2020 | $2.0 \times 10^3$ | July 18, 2020 | $3.0 \times 10^3$ | Fig. S1(d) |
| Open system results | April 3, 2020 | $2.1 \times 10^3$ | July 22, 2020 | $3.4 \times 10^3$ | main text |

Note that, for the closed system setting, the match between *J*(*t*) and the real data is not as good as that in the open system setting shown in the main text, indicating the necessity to model the State of Arizona as an open system. Comparing the results for the closed and open system settings, we see that the largest difference occurs for case (c) where testing is insufficient and control measures are not stringent. In this case, the realistic open system will endure a longer epidemic by about 9 days and an increase of about 1200 in the final confirmed cases. For case (d) where testing is insufficient but control measures are more strict, the differences between the closed and open system settings are much less. These results call for vigorous enforcement of government control measures.

### Supplementary Note 3: Epidemic scenarios in New York City as a closed system

We simulate four distinct epidemic scenarios for New York City as a closed system with different combinations of testing capability and strength of control measures. The City population is about 8.51 millions. The values of the parameters *H*^*^ (0) and *β*^*^ are estimated using the data of daily number of confirmed cases on and before March 29. The results are listed in the table below.

| Parameter values | $T_{inflection}$ | $I_{peak}$ | $T_{ending}$ | $J_{final}$ | Figure |
| --- | --- | --- | --- | --- | --- |
| $\eta = 0.5, \lambda = 0.18$ | April 5, 2020 | $1.1 \times 10^5$ | September 8, 2020 | $1.8 \times 10^5$ | Fig. S2(a) |
| $\eta = 0.5, \lambda = 0.24$ | April 3, 2020 | $0.9 \times 10^5$ | July 8, 2020 | $1.3 \times 10^5$ | Fig. S2(b) |
| $\eta = 0.8, \lambda = 0.18$ | April 6, 2020 | $1.1 \times 10^5$ | October 15, 2020 | $1.9 \times 10^5$ | Fig. S2(c) |
| $\eta = 0.8, \lambda = 0.24$ | April 3, 2020 | $0.9 \times 10^5$ | July 24, 2020 | $1.3 \times 10^5$ | Fig. S2(d) |

A comparison between the projected *J*(*t*) in different panels of Fig. S2 and the available data up to April 12 indicates that Figs. S2(b) and S2(d) represent two likely scenarios for New York City, Note that, in both scenarios, the value of, λ is 0.24, which is consistent with the strict lockdown and restriction measures imposed by the City.

### Supplementary Note 4: Epidemic scenarios in New York State as a closed system

In the main text, the predicted epidemic scenarios for the State of New York obtained using the realistic, dual system, coupled five-state model. To assess the effect of interstate traffic on the epidemic, here we study the hypothetical setting where the State is treated as a closed system. Let *T*_*inflection*_ be the date of occurrence of the inflection point, *I*_*peak*_ be the peaked value of *I*(*t*), *T*_*ending*_ be the ending date of the epidemic, and *J*_*final*_ be the final number of confirmed cases. Four possible epidemic scenarios are shown in Fig. S3. The results, in comparison with those for the corresponding open system in the main text, are listed in the table below.

| Parameter values | $T_{inflection}$ | $I_{peak}$ | $T_{ending}$ | $J_{final}$ | Figure |
| --- | --- | --- | --- | --- | --- |
| $\eta = 0.5, \lambda = 0.18$ | April 8, 2020 | $3.2 \times 10^5$ | November 19, 2020 | $5.8 \times 10^5$ | Fig. S3(a) |
| Open system results | April 8, 2020 | $3.0 \times 10^5$ | November 21, 2020 | $5.5 \times 10^5$ | main text |
| $\eta = 0.5, \lambda = 0.24$ | April 5, 2020 | $2.5 \times 10^5$ | August 2, 2020 | $3.5 \times 10^5$ | Fig. S3(b) |
| Open system results | April 5, 2020 | $2.4 \times 10^5$ | August 1, 2020 | $3.3 \times 10^5$ | main text |
| $\eta = 0.8, \lambda = 0.18$ | April 9, 2020 | $3.6 \times 10^5$ | **January 16, 2021** | $7.6 \times 10^5$ | Fig. S3(c) |
| Open system results | April 9, 2020 | $3.5 \times 10^5$ | **May 30, 2021** | $7.5 \times 10^5$ | main text |
| $\eta = 0.8, \lambda = 0.24$ | April 6, 2020 | $2.8 \times 10^5$ | September 5, 2020 | $4.2 \times 10^5$ | Fig. S3(d) |
| Open system results | April 6, 2020 | $2.7 \times 10^5$ | September 6, 2020 | $4.0 \times 10^5$ | main text |

Comparing the predictions from the closed and open system settings, we see that the main difference occurs in the epidemic ending time for case (c): η = 0.8 and λ = 0.18, where the undocumented infection rate is as high as 80% and the government control measures are not vigorous. In this case, if New York State is treated as a closed system, the epidemic would end on January 16, 2021. However, in the open system setting, the ending date would be more than four months later: May 30, 2021. This means that, if testing and surveillance are largely insufficient, loose control measures would significantly prolong the epidemic due to interstate travel. Indeed, strict implementation of the control measures would diminish the detrimental effects of interstate travel because the predictions from the closed and open system settings are nearly identical even for lack of sufficient testing, as for case (d). These results further highlight the importance of strict implementation and enforcement of government control and prevention measures, corroborating the results in Figs. 3 and 4 in the main text.

### Supplementary Note 5: Epidemic scenarios in USA as a closed system

We treat the US as a closed system, neglecting the influence of input cases from foreign countries after January 30. The total population is *N* = 3.3 × 10^8^. The results of four predicted scenarios are listed in the table below.

| Parameter values | $T_{inflection}$ | $I_{peak}$ | $T_{ending}$ | $J_{final}$ | Figure |
| --- | --- | --- | --- | --- | --- |
| $\eta = 0.5, \lambda = 0.18$ | April 7, 2020 | $9.0 \times 10^5$ | December 14, 2020 | $1.7 \times 10^6$ | Fig. S4(a) |
| $\eta = 0.5, \lambda = 0.24$ | April 4, 2020 | $7.3 \times 10^5$ | August 10, 2020 | $10^6$ | Fig. S4(b) |
| $\eta = 0.8, \lambda = 0.18$ | April 9, 2020 | $1.1 \times 10^6$ | July 21, 2021 | $2.9 \times 10^6$ | Fig. S4(c) |
| $\eta = 0.8, \lambda = 0.24$ | April 6, 2020 | $8.2 \times 10^5$ | September 30, 2020 | $1.3 \times 10^6$ | Fig. S4(d) |

From the insets in different panels of Fig. S4, we note a delay in the disappearance of the H-state population in comparison with the I-state - about seven days for cases (a,b,d) but one month for case (c). This means that, if testing and surveillance are insufficient, the number of confirmed cases reaching zero does not mean the end of the epidemic: there are still a non-negligible number of hidden individuals who carry the virus, are asymptomatic, and can infect others, thereby requiring long term placement of control measures to minimize the rise of a second outbreak. In fact, in Wuhan city (the epicenter in China), the number of confirmed cases has been zero for a few weeks, but the lockdown order was in place until around April 8 and social distancing and isolation orders are still being vigorously enforced.

### Supplementary Note 6: Impacts of interstate and intrastate travel restrictions on eventual epidemic size in New York and Arizona

In the main text, results on the impacts of interstate and intrastate traffic restriction on the epidemic duration in New York and Arizona are presented (Fig. 5). Here we present results on the impacts of such travel restrictions on the final epidemic size in the two states, as shown in Fig. S5. When a reasonable level of testing is assumed (*η* = 0.5), the effects of interstate travel restriction on the epidemic sizes are insignificant [Figs. S5(a) and S5(c)]. However, intrastate travel restriction can have an exponential effect on the epidemic sizes [Figs. S5(b) and S5(d)], similar to its effect on the epidemic duration as described in the main text.

### Supplementary Note 7: Epidemic scenarios in New Jersey as an open system

We simulate four distinct epidemic scenarios for the State of New Jersey treated as an open system with different combinations of testing capability and strength of control measures. The starting date of the epidemic is March 5. The population of the State is *N* = 8.8 × 10^6^. The results are shown in Figs. S6(a-d) and are listed in the table below.

| Parameter values | $T_{inflection}$ | $I_{peak}$ | $T_{ending}$ | $J_{final}$ | Figure |
| --- | --- | --- | --- | --- | --- |
| $\eta = 0.5, \lambda = 0.18$ | April 7, 2020 | $5.4 \times 10^4$ | October 11, 2020 | $10^5$ | Fig. S6(a) |
| $\eta = 0.5, \lambda = 0.24$ | April 4, 2020 | $4.4 \times 10^4$ | July 16, 2020 | $6.0 \times 10^4$ | Fig. S6(b) |
| $\eta = 0.8, \lambda = 0.18$ | April 8, 2020 | $5.7 \times 10^4$ | December 19, 2020 | $1.3 \times 10^5$ | Fig. S6(c) |
| $\eta = 0.8, \lambda = 0.24$ | April 5, 2020 | $4.5 \times 10^4$ | August 14, 2020 | $6.7 \times 10^4$ | Fig. S6(d) |

Comparing the model predicted *J*(*t*) with the real data, we find that Figs. S6(a) and S6(c) represent the two likely scenarios for the State of New Jersey. In both scenarios, the value of λ is 0.18, indicating that the government control measures are not vigorous. The results in Figs. S6(b) and S6(d) indicate that imposing strict government control measures can effectively suppress the epidemic even when testing and surveillance are not sufficient.

### Supplementary Note 8: Epidemic scenarios in California as an open system

We simulate four distinct epidemic scenarios for the State of California as an open system with different combinations of testing capability and strength of control measures. The starting date of the epidemic is February 26. The population of the State is *N* = 39 millions. The results are shown in Figs. S7(a-d) and are listed in the table below.

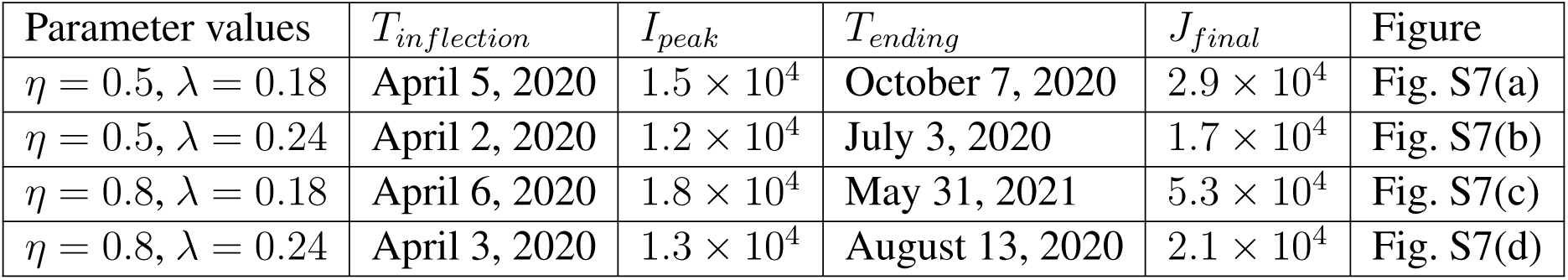

| Parameter values | $T_{inflection}$ | $I_{peak}$ | $T_{ending}$ | $J_{final}$ | Figure |
| --- | --- | --- | --- | --- | --- |
| $\eta = 0.5, \lambda = 0.18$ | April 5, 2020 | $1.5 \times 10^4$ | October 7, 2020 | $2.9 \times 10^4$ | Fig. S7(a) |
| $\eta = 0.5, \lambda = 0.24$ | April 2, 2020 | $1.2 \times 10^4$ | July 3, 2020 | $1.7 \times 10^4$ | Fig. S7(b) |
| $\eta = 0.8, \lambda = 0.18$ | April 6, 2020 | $1.8 \times 10^4$ | May 31, 2021 | $5.3 \times 10^4$ | Fig. S7(c) |
| $\eta = 0.8, \lambda = 0.24$ | April 3, 2020 | $1.3 \times 10^4$ | August 13, 2020 | $2.1 \times 10^4$ | Fig. S7(d) |

Comparing the model predicted *J*(*t*) with the real data, we find that Fig. S7(c) represents a likely scenario for the State of California, which indicates that both government testing services and the control measures are not vigorous enough. Also, comparing Fig. S7(b) with Fig. S7(d) reveals that, when the government testing and surveillance services are significantly reduced, insofar as a stringent set of control measures is in place, the resulting epidemic would be worse but to a relatively small extent, unequivocally pointing at the extremely important role of placing vigorous government control measures in suppressing the epidemic.

### Supplementary Note 9: Epidemic scenarios in Michigan as an open system

We simulate four distinct epidemic scenarios for the State of Michigan as an open system with different combinations of testing capability and strength of control measures. The starting date of the epidemic is March 11. The population of the State is *N* = 9, 986, 857. The results are shown in Figs. S8(a-d) and are listed in the table below.

| Parameter values | $T_{inflection}$ | $I_{peak}$ | $T_{ending}$ | $J_{final}$ | Figure |
| --- | --- | --- | --- | --- | --- |
| $\eta = 0.5, \lambda = 0.18$ | April 9, 2020 | $3.5 \times 10^4$ | November 1, 2020 | $6.7 \times 10^4$ | Fig. S8(a) |
| $\eta = 0.5, \lambda = 0.24$ | April 6, 2020 | $2.8 \times 10^4$ | July 17, 2020 | $3.8 \times 10^4$ | Fig. S8(b) |
| $\eta = 0.8, \lambda = 0.18$ | April 10, 2020 | $3.5 \times 10^4$ | March 23, 2021 | $9.0 \times 10^4$ | Fig. S8(c) |
| $\eta = 0.8, \lambda = 0.24$ | April 7, 2020 | $2.7 \times 10^4$ | August 19, 2020 | $4.0 \times 10^4$ | Fig. S8(d) |

Comparing the model predicted *J*(*t*) with the real data, we find that Fig. S8(d) represents a likely scenario for the State of Michigan. Also, comparing Fig. S8(b) with Fig. S8(d) reveals the important role of placing strict government control measures in suppressing the epidemic.

### Supplementary Note 10: Epidemic scenarios in Florida as an open system

We simulate four distinct epidemic scenarios for the State of Florida as an open system with different combinations of testing capability and strength of control measures. The starting date of the epidemic is March 2. The population of the State is *N* = 21, 477, 737. The results are shown in Figs. S9(a-d) and are listed in the table below.

| Parameter values | $T_{inflection}$ | $I_{peak}$ | $T_{ending}$ | $J_{final}$ | Figure |
| --- | --- | --- | --- | --- | --- |
| $\eta = 0.5, \lambda = 0.18$ | April 9, 2020 | $2.5 \times 10^4$ | November 3, 2020 | $5.0 \times 10^4$ | Fig. S9(a) |
| $\eta = 0.5, \lambda = 0.24$ | April 6, 2020 | $2.0 \times 10^4$ | July 14, 2020 | $2.8 \times 10^4$ | Fig. S9(b) |
| $\eta = 0.8, \lambda = 0.18$ | April 10, 2020 | $2.8 \times 10^4$ | May 8, 2021 | $7.8 \times 10^4$ | Fig. S9(c) |
| $\eta = 0.8, \lambda = 0.24$ | April 7, 2020 | $2.1 \times 10^4$ | August 19, 2020 | $3.2 \times 10^4$ | Fig. S9(d) |

Comparing the model predicted *J*(*t*) with the real data, we find that Figs. S9(b) and S9(d) represent two likely scenarios for the State of Florida.

### Supplementary Note 11: Epidemic scenarios in Illinois as an open system

We simulate four distinct epidemic scenarios for the State of Illinois as an open system with different combinations of testing capability and strength of control measures. The starting date of the epidemic is February 26. The population of the State is *N* = 12, 671, 821. The results are shown in Figs. S10(a-d) and are listed in the table below.

| Parameter values | $T_{inflection}$ | $I_{peak}$ | $T_{ending}$ | $J_{final}$ | Figure |
| --- | --- | --- | --- | --- | --- |
| $\eta = 0.5, \lambda = 0.18$ | April 6, 2020 | $1.8 \times 10^4$ | October 24, 2020 | $3.5 \times 10^4$ | Fig. S10(a) |
| $\eta = 0.5, \lambda = 0.24$ | April 4, 2020 | $1.4 \times 10^4$ | July 9, 2020 | $1.9 \times 10^4$ | Fig. S10(b) |
| $\eta = 0.8, \lambda = 0.18$ | April 8, 2020 | $1.9 \times 10^4$ | April 21, 2021 | $5.4 \times 10^4$ | Fig. S10(c) |
| $\eta = 0.8, \lambda = 0.24$ | April 5, 2020 | $1.4 \times 10^4$ | August 14, 2020 | $2.2 \times 10^4$ | Fig. S10(d) |

Comparing the model predicted *J*(*t*) with the real data, we find that Figs. S10(a) and S10(c) represent two likely scenarios for the State of Illinois.

### Supplementary Note 12: Epidemic scenarios in Massachusetts as an open system

We simulate four distinct epidemic scenarios for the State of Massachusetts as an open system with different combinations of testing capability and strength of control measures. The starting date of the epidemic is March 2. The population of the State is *N* = 6, 892, 503. The results are shown in Figs. S11(a-d) and are listed in the table below.

| Parameter values | $T_{inflection}$ | $I_{peak}$ | $T_{ending}$ | $J_{final}$ | Figure |
| --- | --- | --- | --- | --- | --- |
| $\eta = 0.5, \lambda = 0.18$ | April 10, 2020 | $3.7 \times 10^4$ | October 30, 2020 | $7.0 \times 10^4$ | Fig. S11(a) |
| $\eta = 0.5, \lambda = 0.24$ | April 7, 2020 | $2.9 \times 10^4$ | July 17, 2020 | $4.0 \times 10^4$ | Fig. S11(b) |
| $\eta = 0.8, \lambda = 0.18$ | April 11, 2020 | $3.6 \times 10^4$ | March 25, 2021 | $9.0 \times 10^4$ | Fig. S11(c) |
| $\eta = 0.8, \lambda = 0.24$ | April 8, 2020 | $2.7 \times 10^4$ | August 19, 2020 | $4.1 \times 10^4$ | Fig. S11(d) |

Comparing the model predicted *J*(*t*) with the real data, we find that Fig. S11(d) represents a likely scenario for the State of Massachusetts, suggesting that the current government control measures are stringent.

### Supplementary Note 13: Epidemic scenarios in Louisiana as an open system

We simulate four distinct epidemic scenarios for the State of Louisiana as an open system with different combinations of testing capability and strength of control measures. The starting date of the epidemic is March 9. The population of the State is *N* = 4, 648, 794. The results are shown in Figs. S12(a-d) and are listed in the table below.

| Parameter values | $T_{inflection}$ | $I_{peak}$ | $T_{ending}$ | $J_{final}$ | Figure |
| --- | --- | --- | --- | --- | --- |
| $\eta = 0.5, \lambda = 0.18$ | April 8, 2020 | $2.3 \times 10^4$ | October 4, 2020 | $4.2 \times 10^4$ | Fig. S12(a) |
| $\eta = 0.5, \lambda = 0.24$ | April 5, 2020 | $1.9 \times 10^4$ | July 8, 2020 | $2.6 \times 10^4$ | Fig. S12(b) |
| $\eta = 0.8, \lambda = 0.18$ | April 10, 2020 | $2.6 \times 10^4$ | February 24, 2021 | $6.3 \times 10^4$ | Fig. S12(c) |
| $\eta = 0.8, \lambda = 0.24$ | April 7, 2020 | $2.0 \times 10^4$ | August 13, 2020 | $2.9 \times 10^4$ | Fig. S12(d) |

Comparing the model predicted *J*(*t*) with the real data, we find that Figs. S12(b) and S12(d) represent two likely scenarios for the State of Louisiana.

### Supplementary Note 14: Uncertainty ranges of selected predictions

Due to the uncertainties in the estimated model parameters, for each scenario, the predicted outcomes have a range. To determine the daily ranges for all cases is computationally prohibitive. For the purpose of illustration, we present the range of predicted quantities for three examples: New York and Arizona as an open system and the US as a closed system.

A difficulty in assessing the uncertainties in the estimated parameter values is the lack of sufficient samples or data. Our solution is to resort to the non-parametric bootstrap method [24] and apply it to *J*(*t*). The basic assumption is that the data point *J*(*t*_*i*_) obeys a normal distribution. The parameter uncertainties can be obtained through multiple sampling of the optimal model. In particular, from the data of confirmed infections *J*(*t*_1_),*J* (*t*_2_),…, *J*(*t*_*n*_), we first use a least squares fitting to obtain the optimal estimates of the parameters: 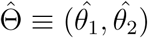, where 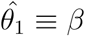 and 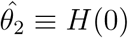. Let 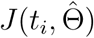 denote the model predicted data set of confirmed infections from the estimated parameter values 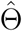. For the normal distribution of 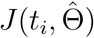 at time *t*_*i*_, its mean is taken to be the actual data point *J*(*t*_*i*_) and its standard deviation is approximated by that of the increments in the number of confirmed cases within five days. We then generate *S* = 10^3^ realizations of the data set 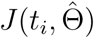, denoted as 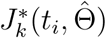 for *t* = 1,…, *n* and *k* = 1,…, *S*. For each data realization, the optimal parameter values can be estimated, generating the corresponding prediction results. Repeating this procedure for each of the *S* data sets gives the prediction ranges for the epidemic quantities of interest.

The results with the prediction ranges for the three examples are shown in Figs. S13, S14, and S15. We observe that the uncertainty ranges of the predicted quantities increase with time, as expected. The open system model generates predictions with relatively less uncertainty as compared to those from the closed system model.

**Supplementary Table 1.**
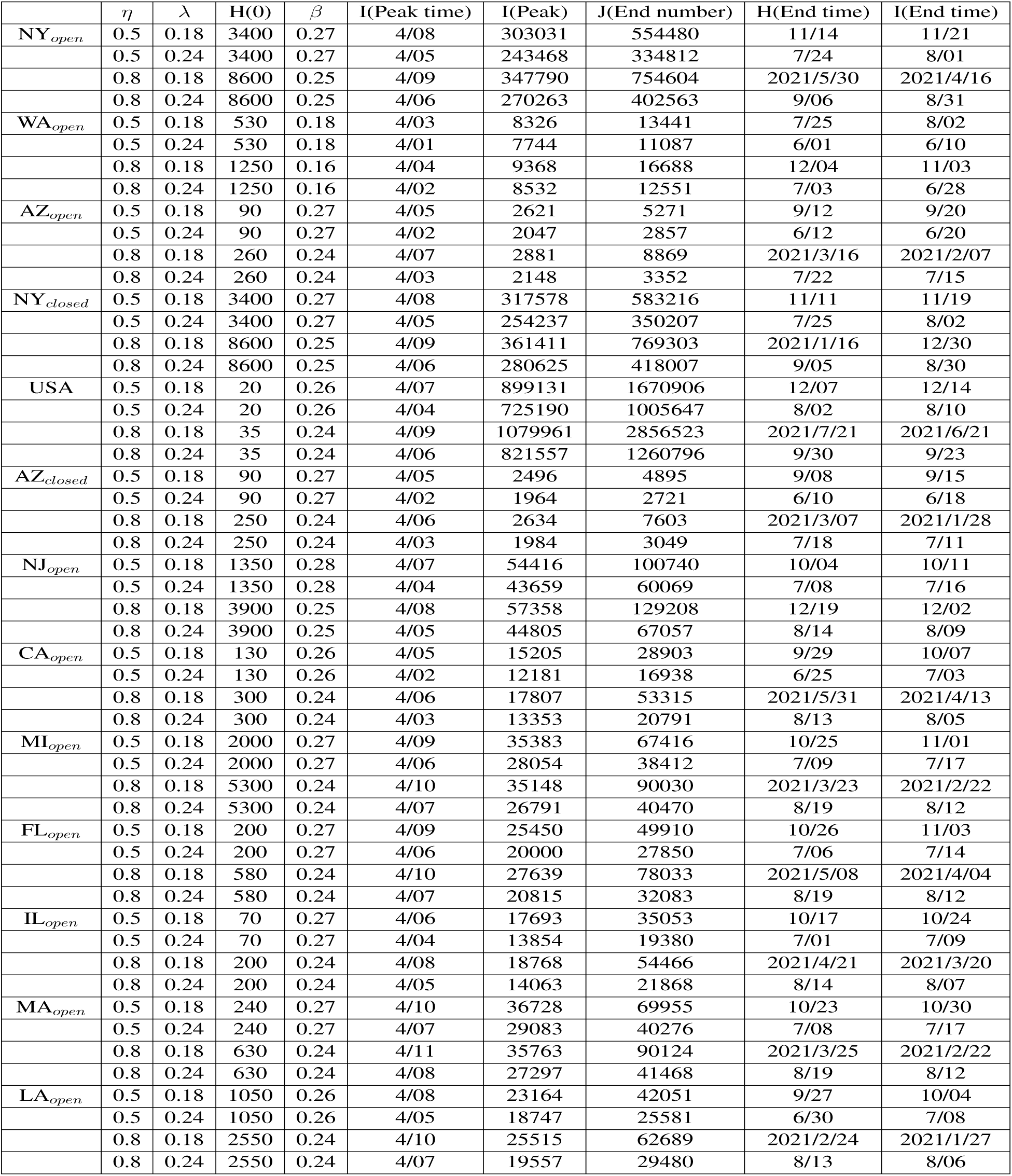
Summary of prediction results for ten States (each as an open system), for New York and Arizona (each as a closed system), and for USA as a closed system.

**Supplementary Table 2.** The dates on which travel restriction orders were issued in the ten States studied. For each state, the date is effectively one on which an exponential decay in intrastate traffic begins.

| State | Date |
| --- | --- |
| Arizona | March 19, 2020 |
| Washington | March 23, 2020 |
| New York | March 22, 2020 |
| New Jersey | March 21, 2020 |
| California | March 19, 2020 |
| Michigan | March 23, 2020 |
| Florida | March 23, 2020 |
| Illinois | March 21, 2020 |
| Massachusetts | March 24, 2020 |
| Louisiana | March 23, 2020 |

**Supplementary Table 3.** Average daily outbound and inbound populations and the total population for the ten States studied

| State | Outbound | Inbound | Population |
| --- | --- | --- | --- |
| Arizona | 13,679 | 6,515 | 7,278,717 |
| Washington | 27,339 | 15,495 | 7,614,893 |
| New York | 55,012 | 133,095 | 19,453,561 |
| New Jersey | 133,507 | 71,056 | 8,882,190 |
| California | 21,171 | 19,979 | 39,512,223 |
| Michigan | 21,179 | 11,625 | 9,986,857 |
| Florida | 28,871 | 22,111 | 21,477,737 |
| Illinois | 48,160 | 45,451 | 12,671,821 |
| Massachusetts | 30,217 | 44,987 | 6,892,503 |
| Louisiana | 11,754 | 14,994 | 4,648,794 |

## Supplementary Figures

**Supplementary Figure S1.**
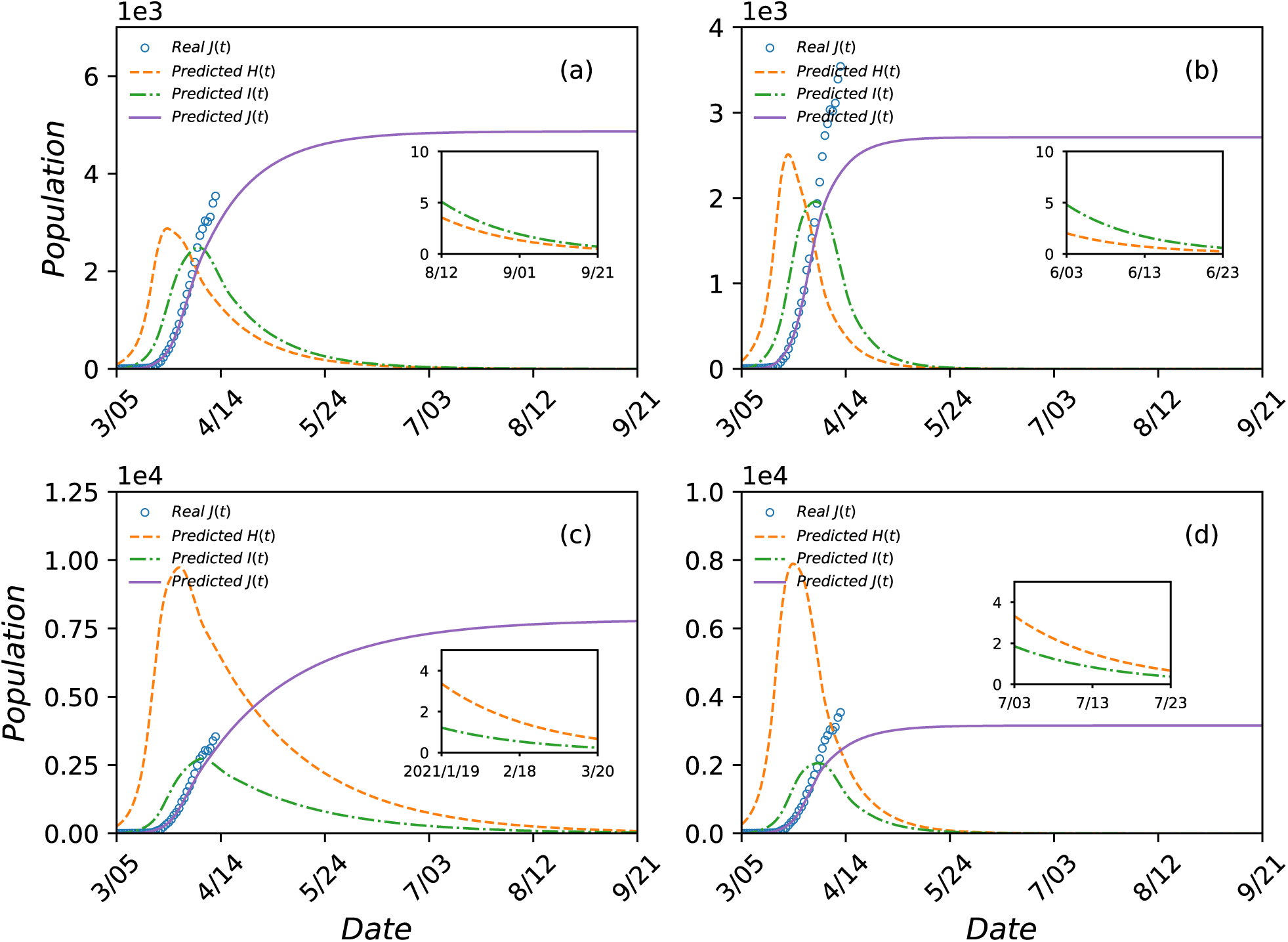
Prediction of COVID-19 epidemic scenarios for the State of Arizona treated as a closed system. Shown are *H*(*t*) (dashed orange curve), *I*(*t*) (green dot-dashed curve), and *J*(*t*) (solid purple curve) from March 5 to September 21 as predicted by the closed five-state model. The open blue circles are the actual data of *J*(*t*) available up to the time of writing. The parameter settings are: (a) *η* = 0.5 and, λ = 0.18, (b) *η* = 0.5 and, λ = 0.24, (c) *η* = 0.8 and, λ = 0.18, and (d) *η* = 0.8 and, λ = 0.24. For (a) and (b), the optimal estimates of the number of initial hidden population and the inflection rate are *H*^*^ (0) = 90 and *β*^*^ = 0.27. For (c) and (d), these values are *H*^*^ (0) = 250 and *β*^*^ = 0.24. The inset in each panel shows the diminishing trend of *H*(*t*) and *I*(*t*) during the last stage of the epidemic.

**Supplementary Figure S2.**
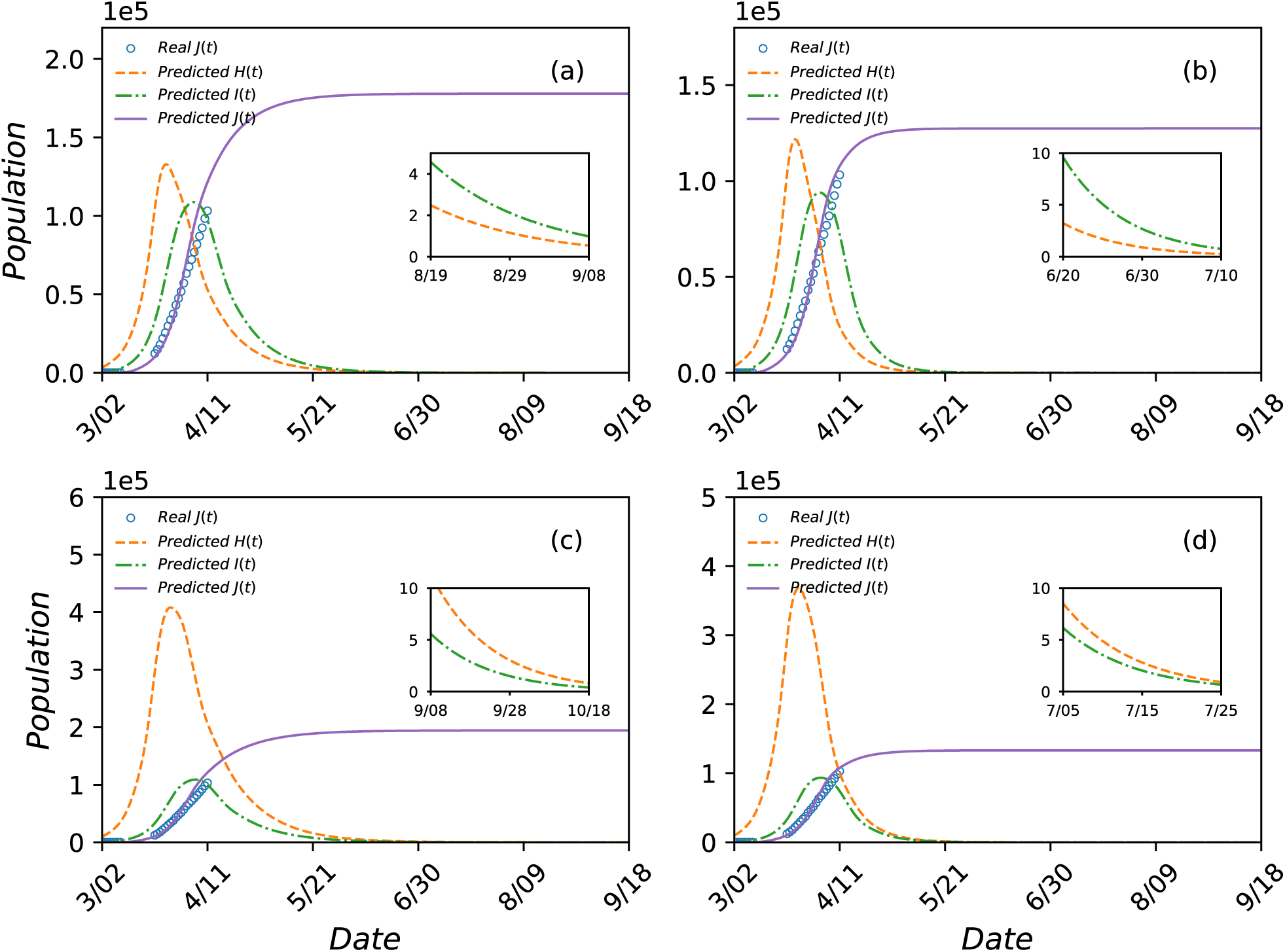
Prediction of COVID-19 epidemic scenarios for the City of New York treated as a closed system. The legends are the same as those in Fig. S1. Shown are the prediction results from the closed five-state model for the time period from March 2 to September 18. The open blue circles are the actual data of *J*(*t*) available up to the time of writing. The parameter settings are: (a) *η* = 0.5 and λ. = 0.18, (b) *η* = 0.5 and λ. = 0.24, (c) *η* = 0.8 and λ = 0.18, and (d) *η* = 0.8 and λ = 0.24. For (a) and (b), the optimal estimates of the number of initial hidden population and the inflection rate are *H*\*(0) = 3400 and *β*^*^ = 0.23. For (c) and (d), these values are *H*^*^ (0) = 10000 and *β*^*^ = 0.2. The inset in each panel shows the diminishing trend of *H*(*t*) and *I*(*t*) during the last stage of the epidemic.

**Supplementary Figure S3.**
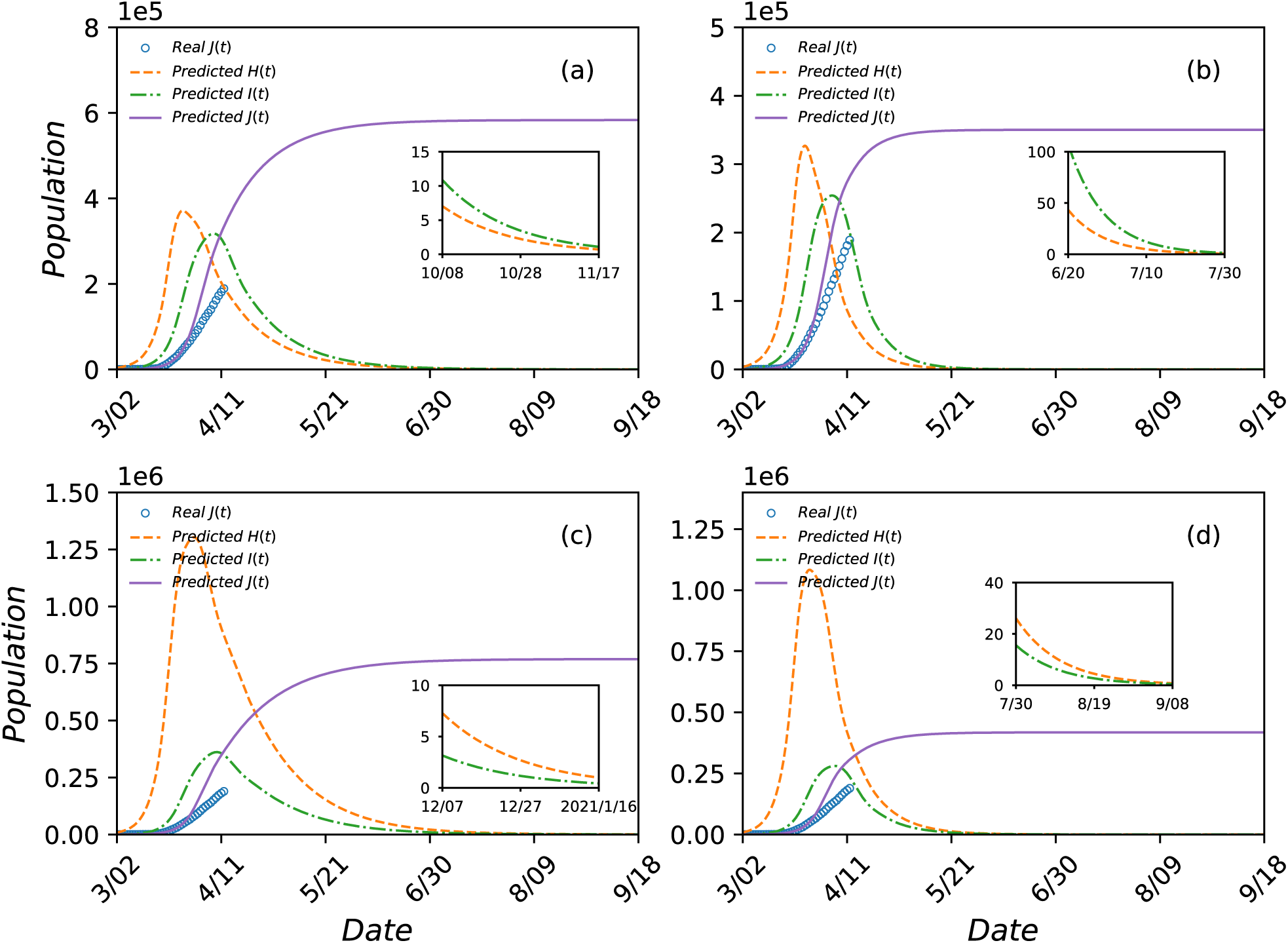
Prediction of COVID-19 epidemic scenarios in New York State as a closed system. The legends are the same as those in Fig. S1. Shown are the prediction results from the closed five-state model. The parameter values are: (a) *η* = 0.5 and λ= 0.18, (b) *η* = 0.5 and λ= 0.24, (c) *η* = 0.8 and λ= 0.18, and (d) *η* = 0.8 and λ= 0.24. For (a) and (b), the optimal estimates of the number of initial hidden population and the inflection rate are *H*^*^ (0) = 3400 and *β*^*^ = 0.27. For (c) and (d), these values are *H*^*^ (0) = 8600 and *β*^*^ = 0.25. The inset in each panel shows the diminishing trend of *H*(*t*) and *I*(*t*) during the last stage of the epidemic.

**Supplementary Figure S4.**
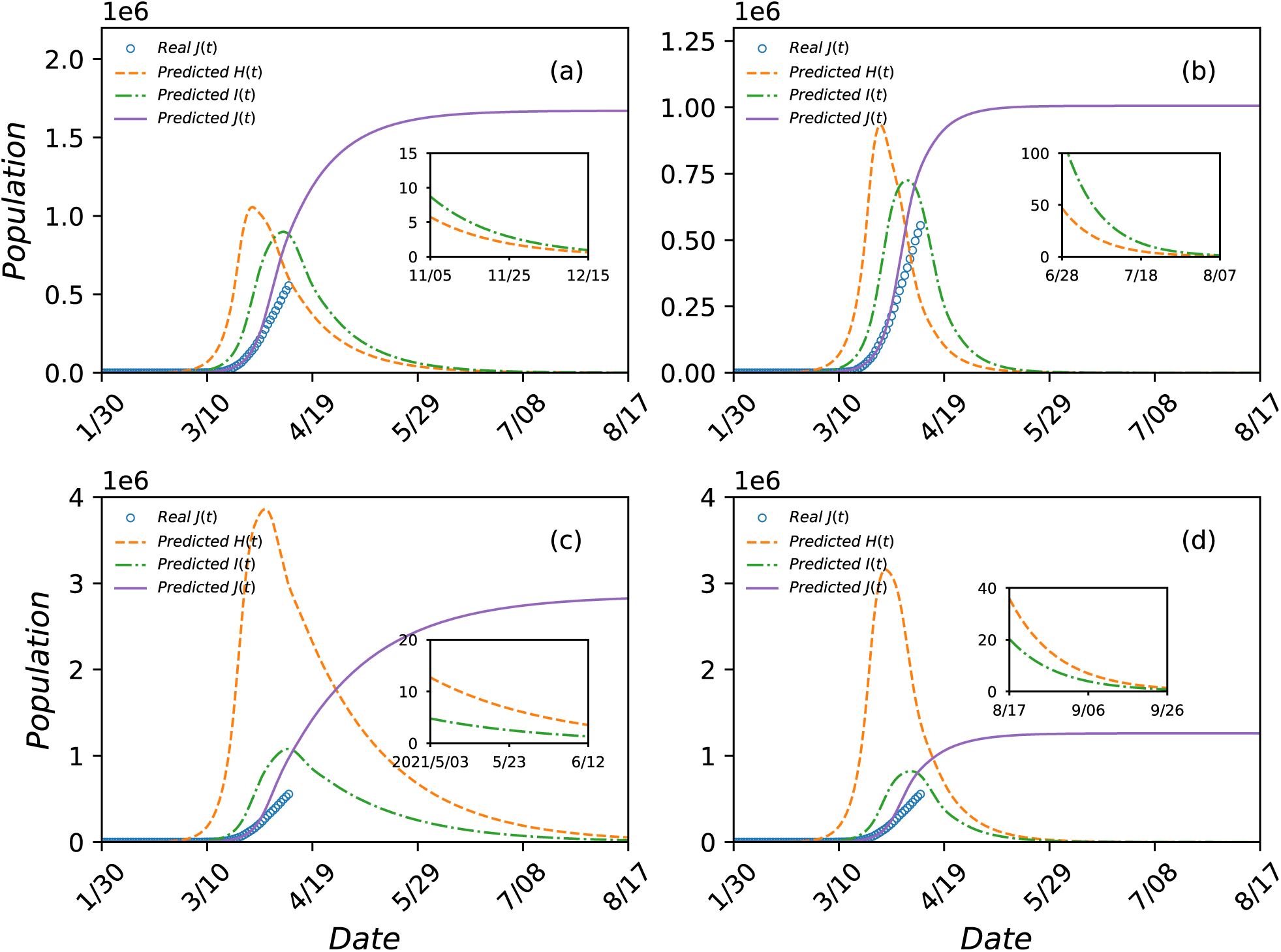
Prediction of COVID-19 epidemic scenarios for USA. The legends are the same as those in Fig. S1. Shown are the prediction results from the closed five-state model for the time period between January 20 to August 17. The open blue circles are the actual data of *J*(*t*) available up to the time of writing. The parameter settings are: (a) *η* = 0.5 and λ= 0.18, (b) *η* = 0.5 and λ= 0.24, (c) *η* = 0.8 and λ= 0.18, and (d) *η* = 0.8 and λ= 0.24. For (a) and (b), the optimal estimates of the number of initial hidden population and the inflection rate are *H*^*^ (0) = 20 and *β*\* = 0.26. For (c) and (d), these values are *H*^*^ (0) = 35 and *β*\* = 0.24. The inset in each panel shows the diminishing trend of *H*(*t*) and *I*(*t*) during the last stage of the epidemic.

**Supplementary Figure S5.**
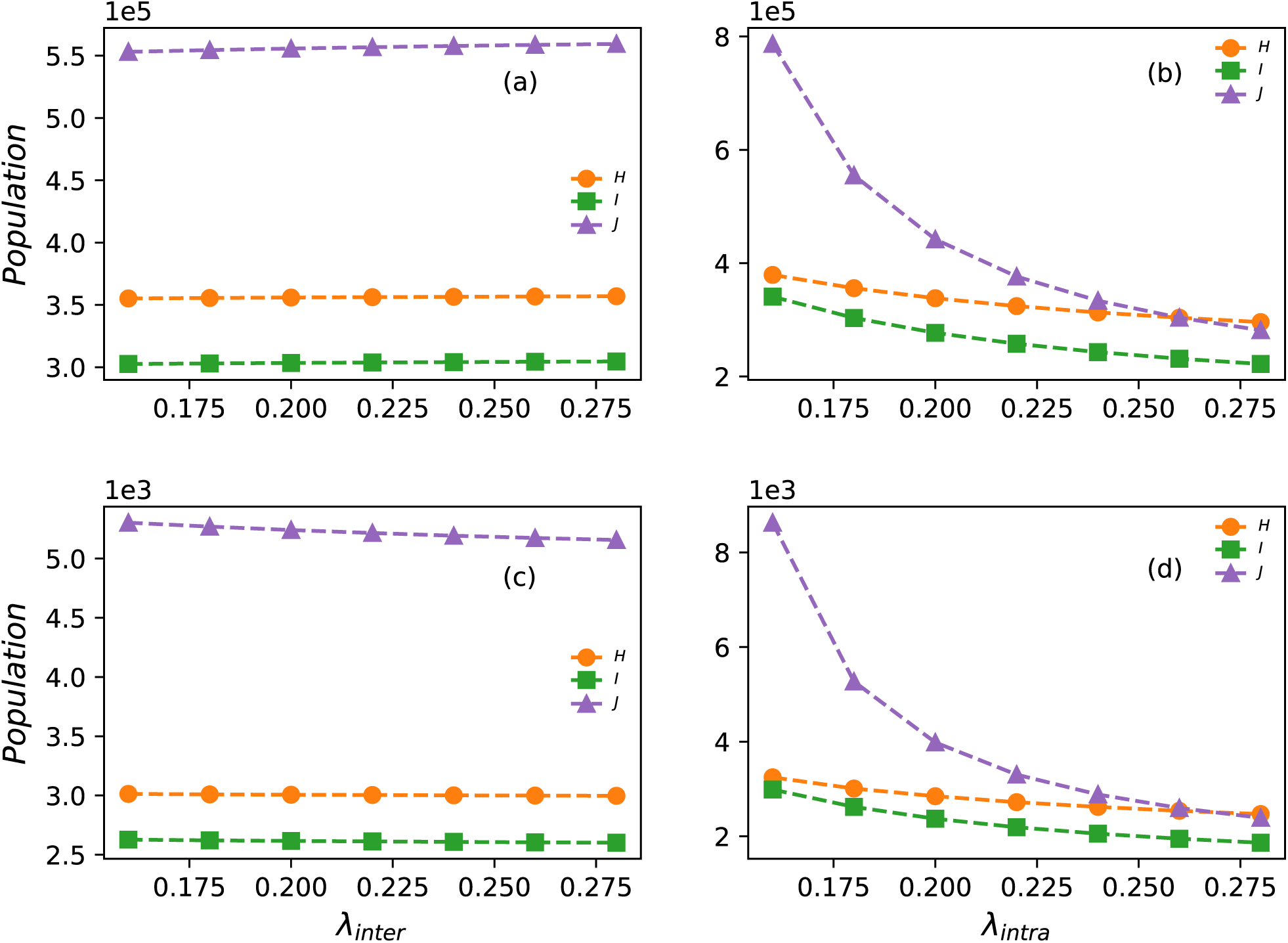
Impact of interstate and intrastate traffic restriction on epidemic size in New York and Arizona. (a) For New York, the final epidemic sizes of H (orange), I (green) and J (purple) versus the interstate traffic reduction rate λ_*inter*_ for λ_*intra*_ = 0.18. (b) For New York, the final sizes versus the intrastate traffic reduction rate λ_*intra*_ for λ_*inter*_ = 0.18. In (a,b), the fraction of undocumented inflections is *η* = 0.5 and the optimized model parameter values are *H*^*^ (0) = 3400 and *β*^*^ = 0.27. (c,d) The corresponding results for Arizona for *η* = 0.5, *H*^*^ (0) = 90, and *β*^*^ = 0.27. For both States, the effects of interstate traffic restriction are relatively small (a,c), but the intrastate traffic restriction leads to an exponential decrease in the final epidemic sizes.

**Supplementary Figure S6.**
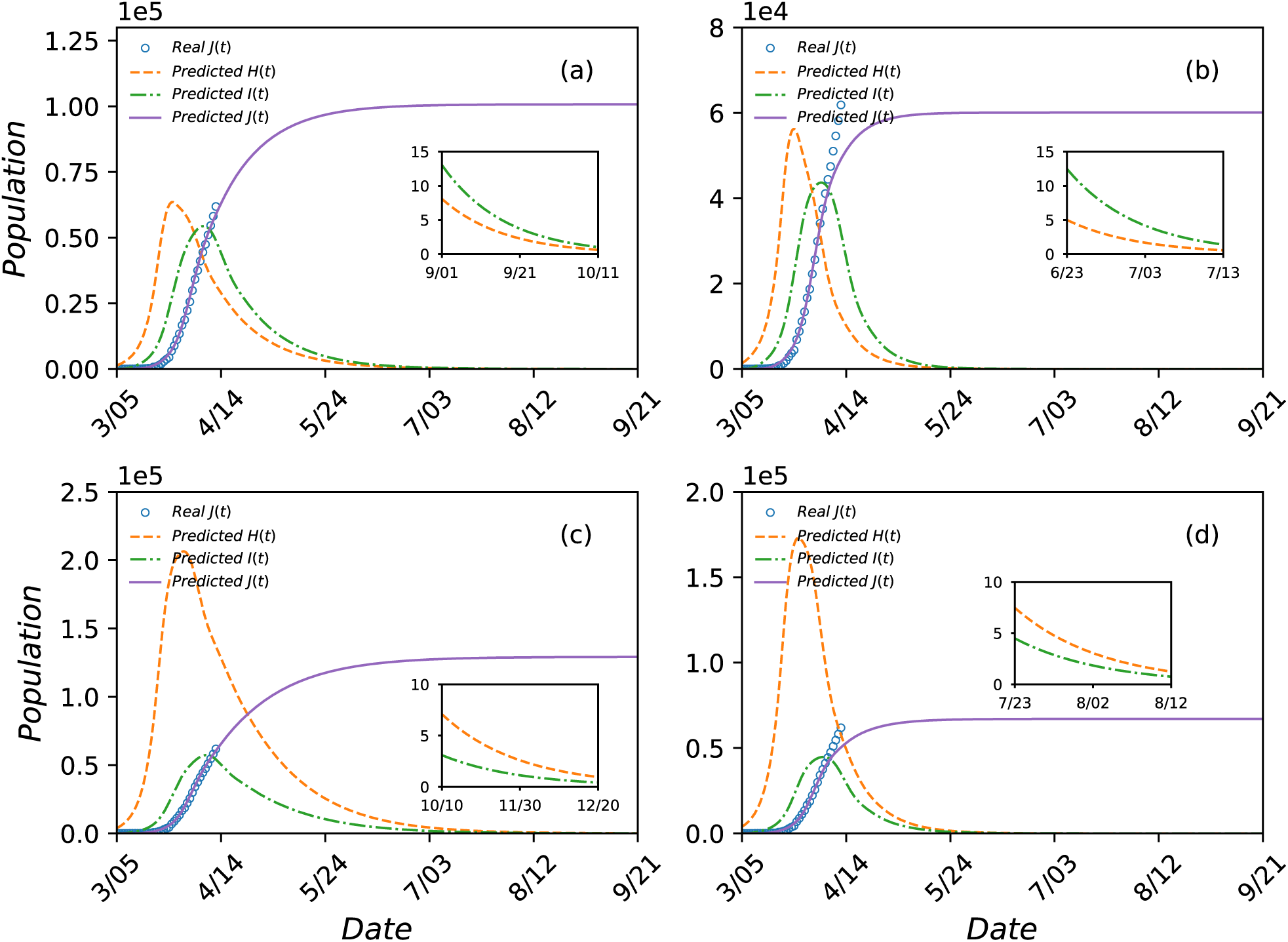
Prediction of COVID-19 epidemic scenarios for the State of New Jersey as an open system. The legends are the same as those in Fig. S1. Shown are the prediction results for the time period from March 5 to September 21. The open blue circles are the actual data of *J*(*t*) available up to the time of writing. The parameter settings are: (a) *η* = 0.5 and λ = 0.18, (b) *η* = 0.5 and λ = 0.24, (c) *η* = 0.8 and λ = 0.18, and (d) *η* = 0.8 and λ = 0.24. For (a) and (b), the optimal estimates of the number of initial hidden population and the inflection rate are *H*^*^ (0) = 1350 and *β*^*^ = 0.28. For (c) and (d), these values are *H*^*^ (0) = 3900 and *β*^*^ = 0.25. The inset in each panel shows the diminishing trend of *H*(*t*) and *I*(*t*) during the last stage of the epidemic.

**Supplementary Figure S7.**
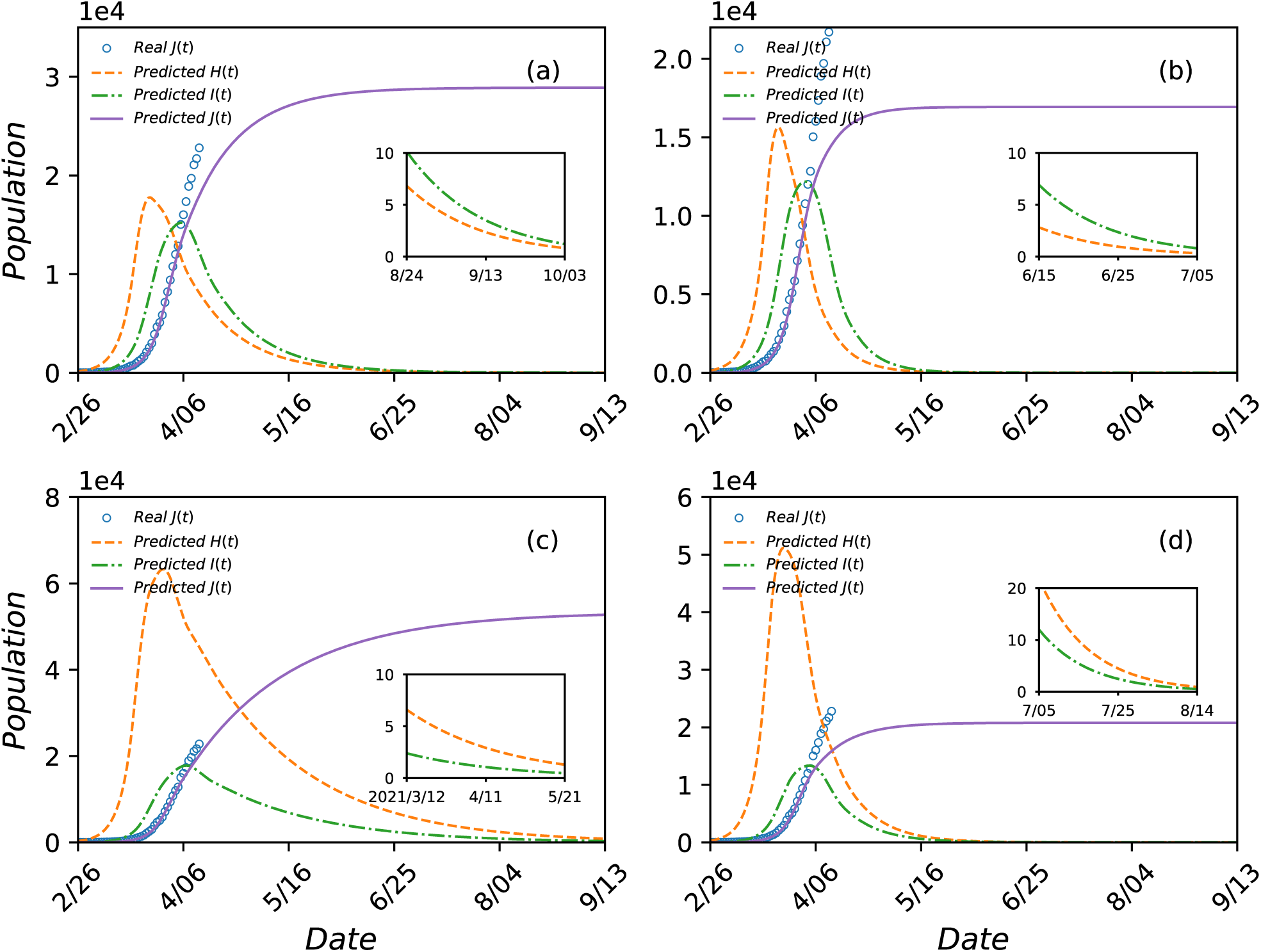
Predicted COVID-19 epidemic scenarios in California as an open system. Legends are the same as those in Fig. S1. The time period covered is from February 26 to September 13, 2020. The parameter settings are: (a) *η* = 0.5 and λ= 0.18, (b) *η* = 0.5 and λ= 0.24, (c) *η* = 0.8 and λ= 0.18, and (d) *η* = 0.8 and λ= 0.24. For (a) and (b), optimization of the model equations gives *H*\*(0) = 130 and *β*^*^ = 0.26. For (c) and (d), the corresponding values are *H*\*(0) = 300 and *β*^*^ = 0.24. The inset in each panel shows the predicted behaviors of *H*(*t*) and *I*(*t*) towards the end of the epidemic.

**Supplementary Figure S8.**
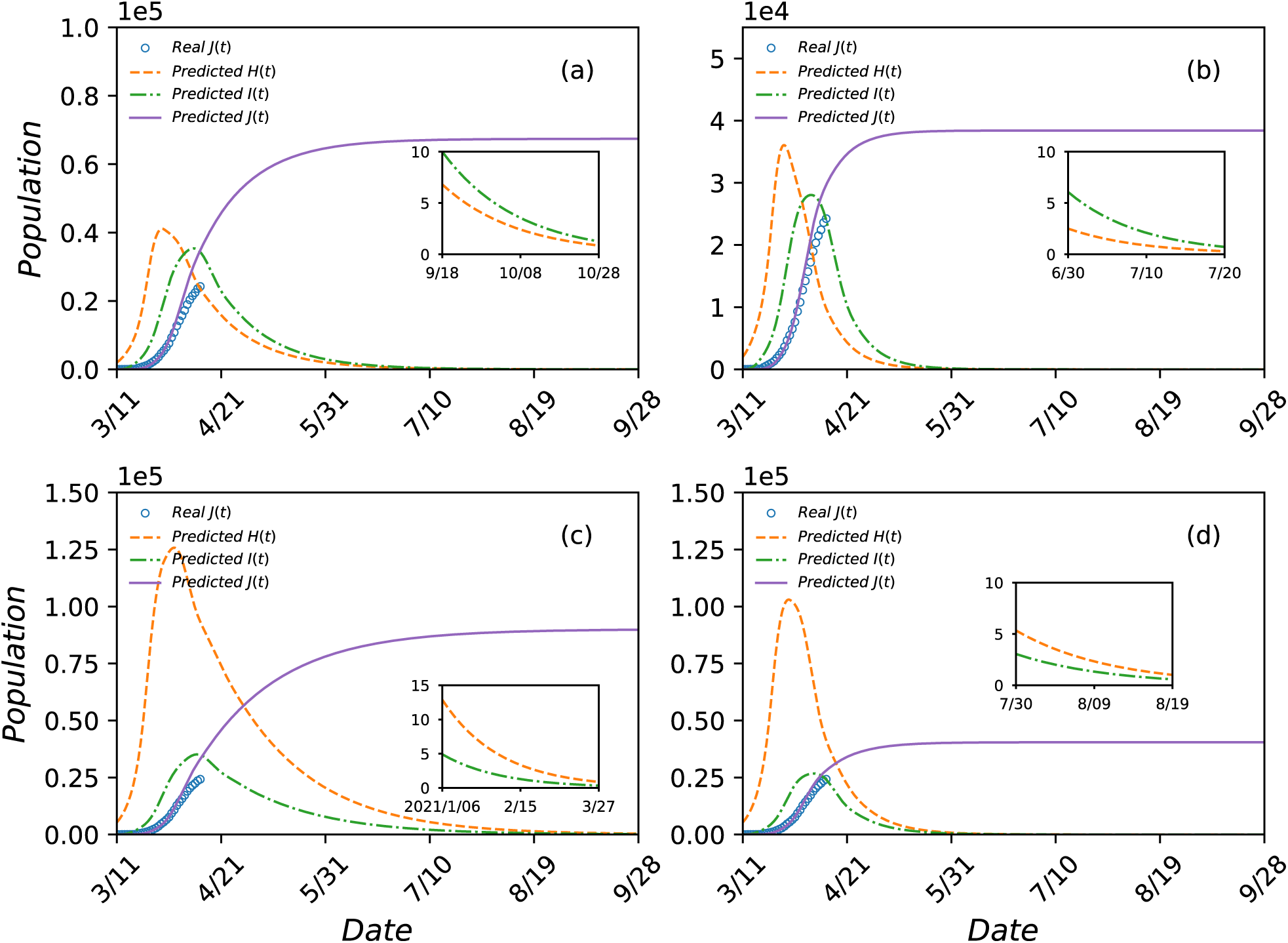
Predicted COVID-19 epidemic scenarios in Michigan as an open system. Legends are the same as those in Fig. S1. The time period covered is from March 11 to September 28, 2020. The parameter settings are: (a) *η* = 0.5 and λ= 0.18, (b) *η* = 0.5 and λ= 0.24, (c) *η* = 0.8 and λ= 0.18, and (d) *η* = 0.8 and λ= 0.24. For (a) and (b), optimization of the model equations gives *H*^*^ (0) = 2000 and *β* ^*^ = 0.27. For (c) and (d), the corresponding values are *H*^*^ (0) = 5300 and *β* ^*^ = 0.24. The inset in each panel shows the predicted behaviors of *H*(*t*) and *I*(*t*) towards the end of the epidemic.

**Supplementary Figure S9.**
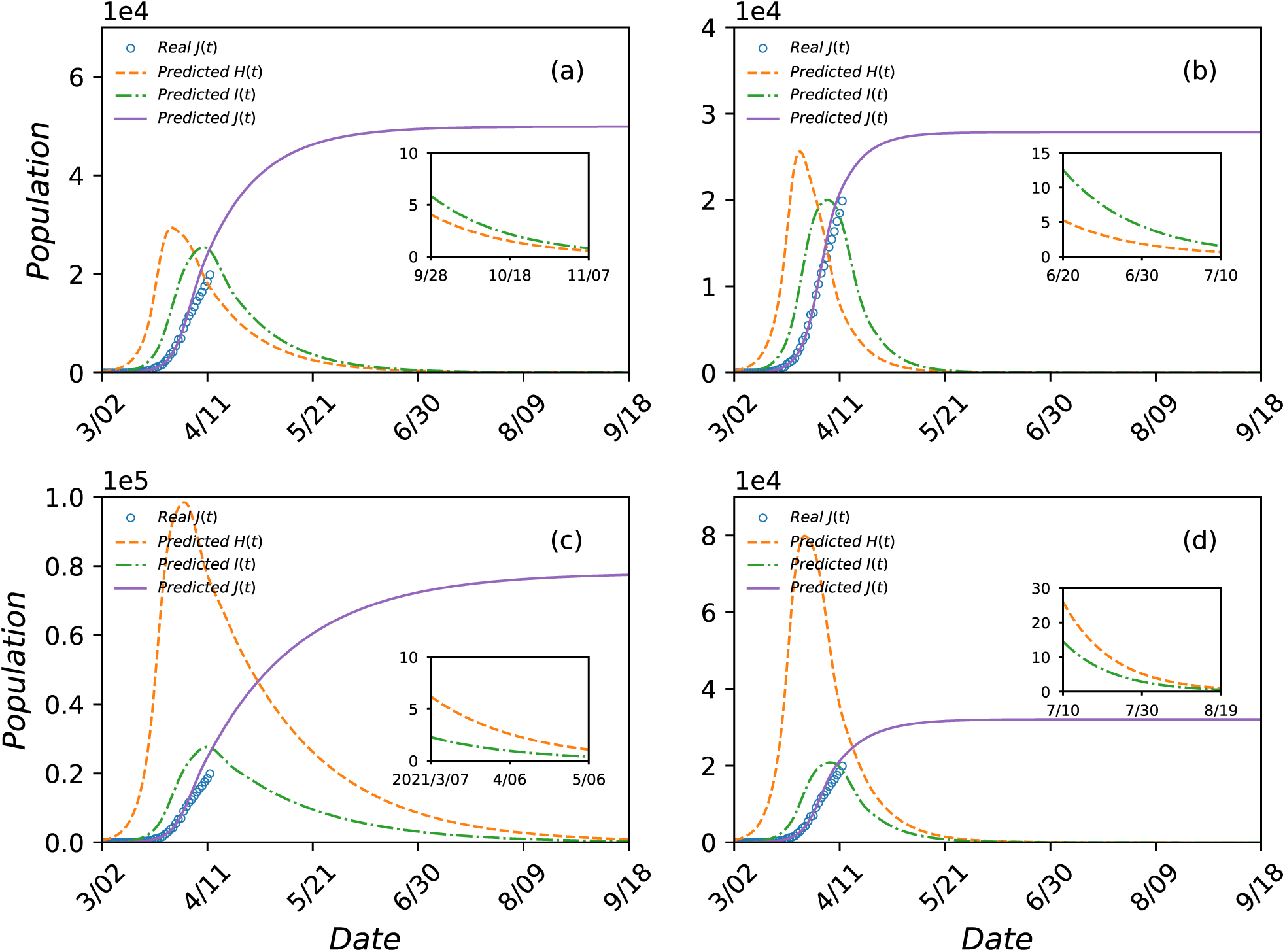
Predicted COVID-19 epidemic scenarios in Florida as an open system. Legends are the same as those in Fig. S3. The time period covered is from March 2 to September 18, 2020. The parameter settings are: (a) *η* = 0.5 and λ= 0.18, (b) *η* = 0.5 and λ= 0.24, (c) *η* = 0.8 and λ= 0.18, and (d) *η* = 0.8 and λ= 0.24. For (a) and (b), optimization of the model equations gives *H*\*(0) = 200 and *β*^*^ = 0.27. For (c) and (d), the corresponding values are *H*\*(0) = 580 and *β*^*^ = 0.24. The inset in each panel shows the predicted behaviors of *H*(*t*) and *I*(*t*) towards the end of the epidemic.

**Supplementary Figure S10.**
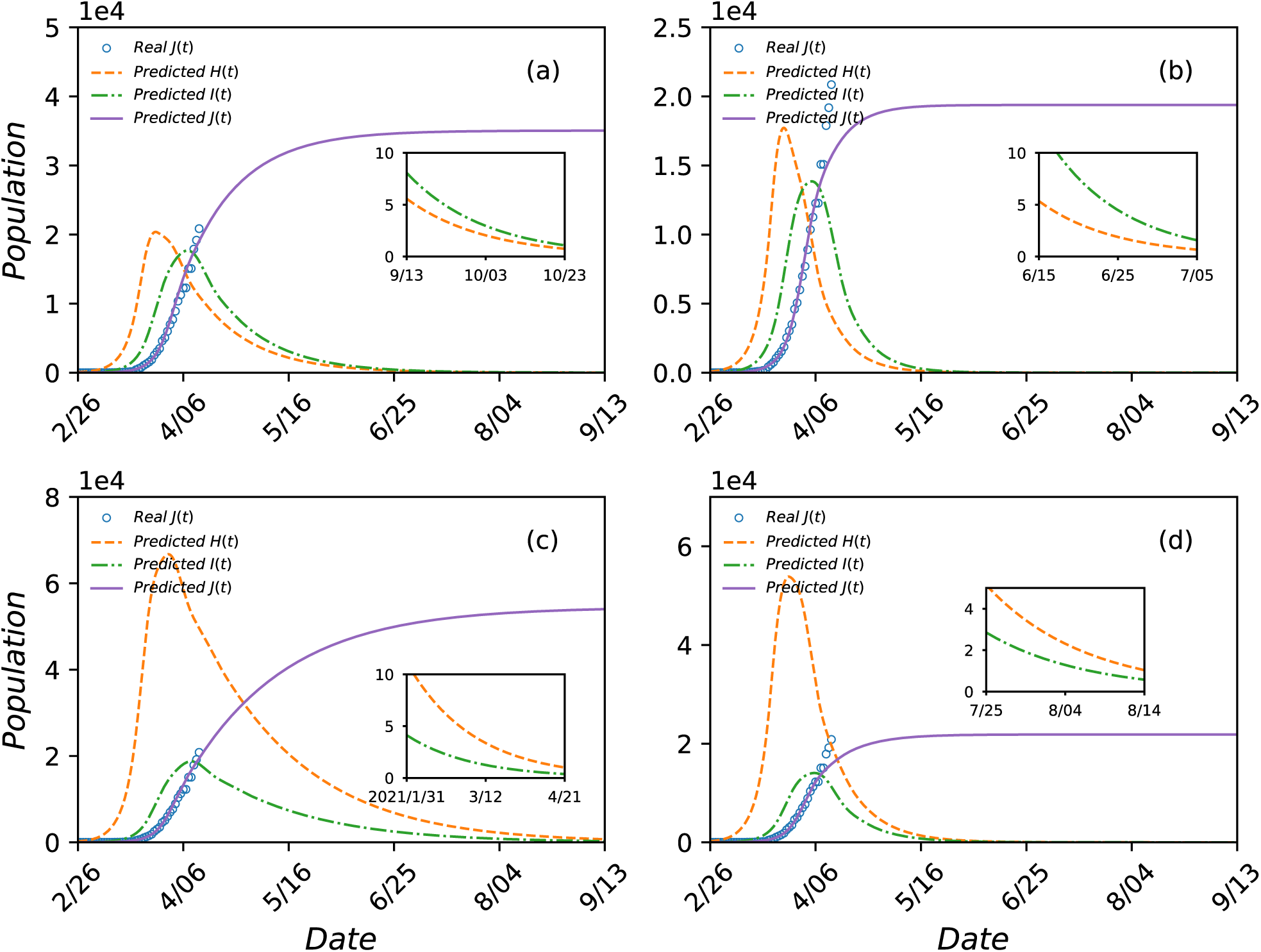
Predicted COVID-19 epidemic scenarios in Illinois as an open system. Legends are the same as those in Fig. S3. The time period covered is from February 26 to September 13, 2020. The parameter settings are (a) *η* = 0.5 and λ= 0.18, (b) *η* = 0.5 and λ= 0.24, (c) *η* = 0.8 and λ= 0.18, and (d) *η* = 0.8 and λ= 0.24. For (a) and (b), optimization of the model equations gives *H*^*^ (0) = 70 and *β*^*^ = 0.27. For (c) and (d), the corresponding values are *H*\*(0) = 200 and *β*^*^ = 0.24. The inset in each panel shows the predicted behaviors of *H*(*t*) and *I*(*t*) towards the end of the epidemic.

**Supplementary Figure S11.**
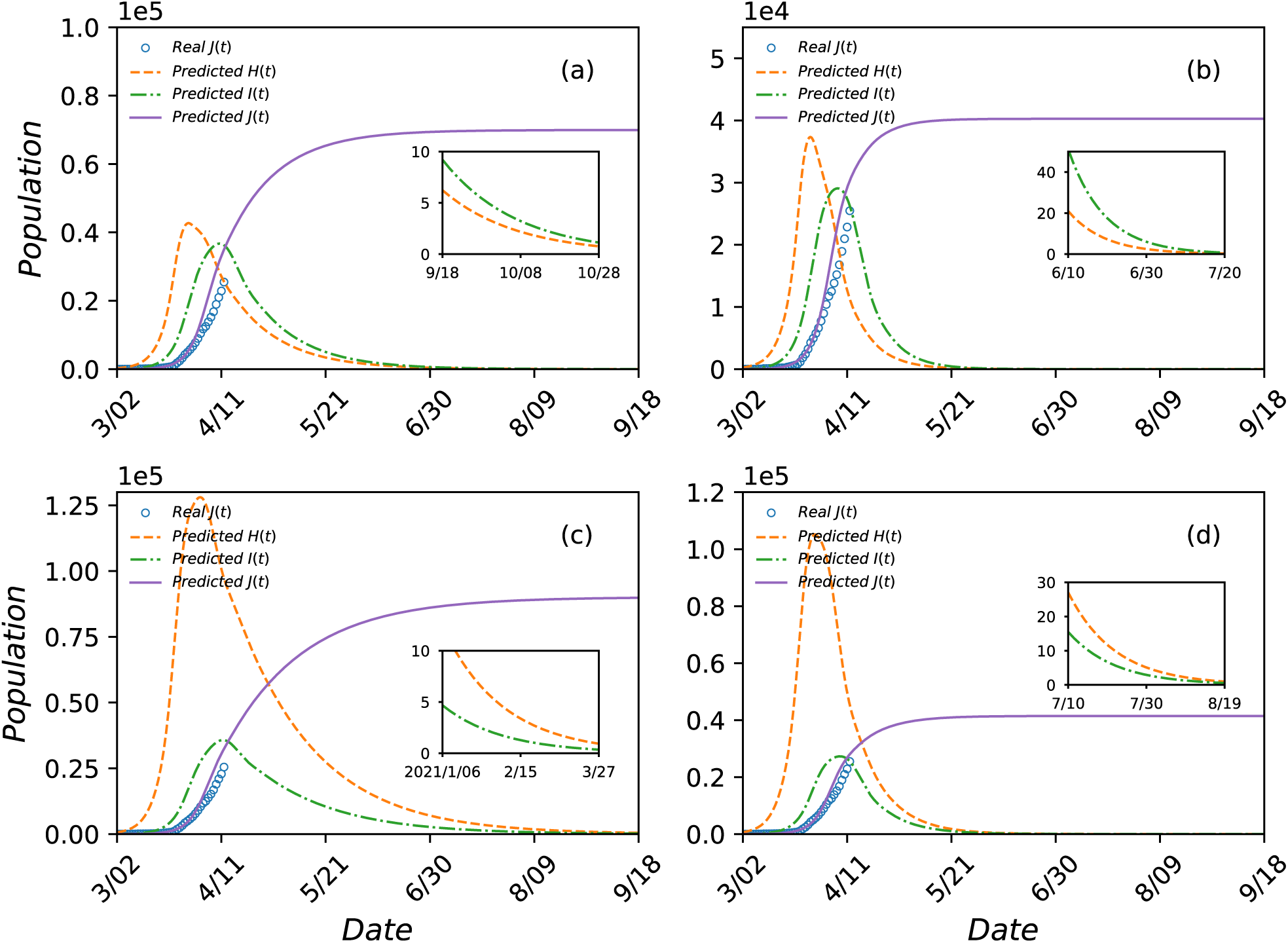
Predicted COVID-19 epidemic scenarios in Massachusetts as an open system. Legends are the same as those in Fig. S3. The time period covered is from March 2 to September 18, 2020. The parameter settings are (a) *η* = 0.5 and λ= 0.18, (b) *η* = 0.5 and λ= 0.24, (c) *η* = 0.8 and λ= 0.18, and (d) *η* = 0.8 and λ= 0.24. For (a) and (b), optimization of the model equations gives *H*^*^ (0) = 240 and *β* ^*^ = 0.27. For (c) and (d), the corresponding values are *H*^*^ (0) = 600 and *β* ^*^ = 0.24. The inset in each panel shows the predicted behaviors of *H*(*t*) and *I*(*t*) towards the end of the epidemic.

**Supplementary Figure S12.**
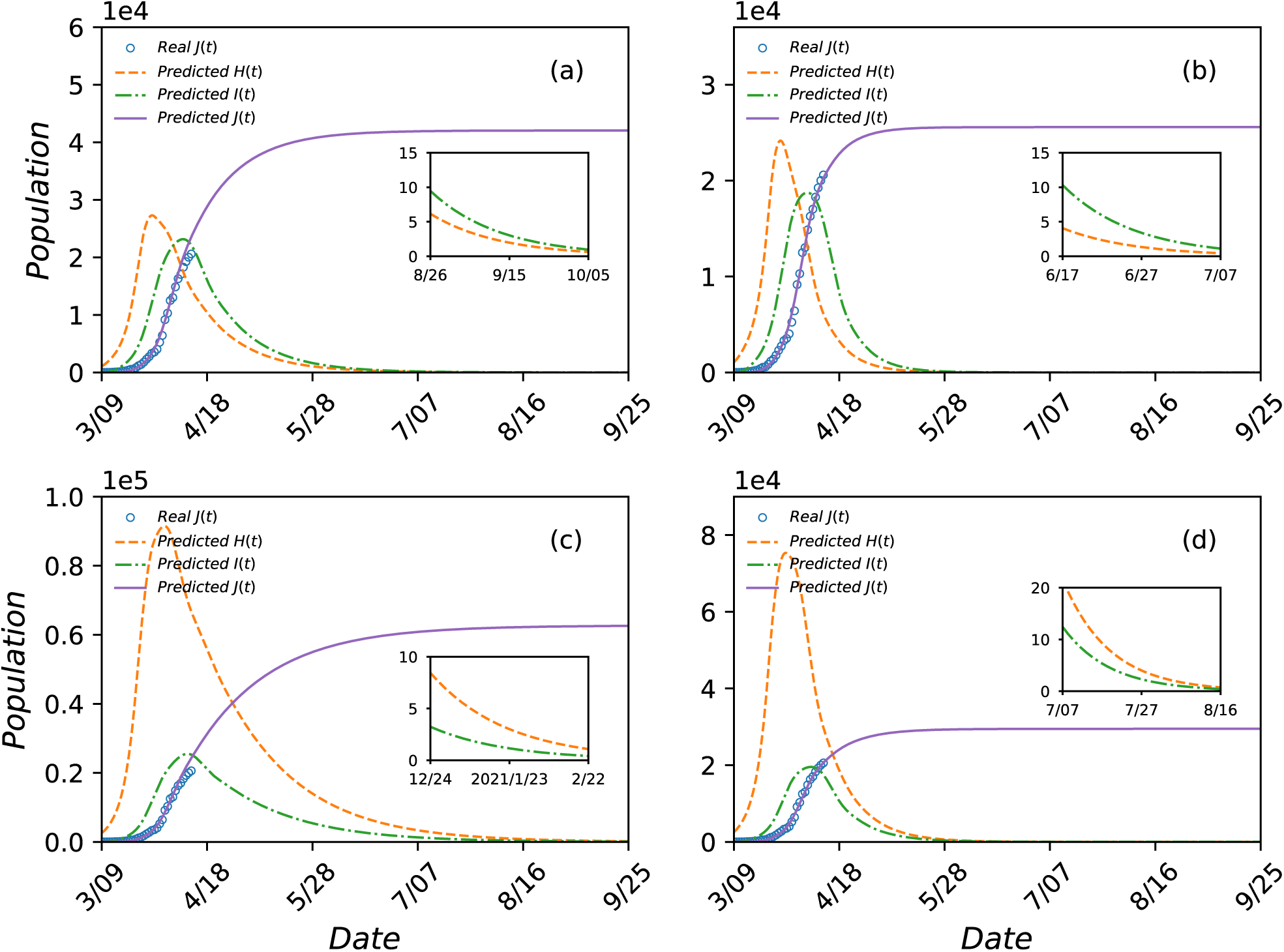
Predicted COVID-19 epidemic scenarios in Louisiana as an open system. Legends are the same as those in Fig. S3. The time period covered is from March 9 to September 25, 2020. The parameter settings are (a) *η* = 0.5 and λ= 0.18, (b) *η* = 0.5 and λ= 0.24, (c) *η* = 0.8 and λ= 0.18, and (d) *η* = 0.8 and λ= 0.24. For (a) and (b), optimization of the model equations gives *H*\*(0) = 1050 and *β*^*^ = 0.26. For (c) and (d), the corresponding values are *H*\*(0) = 2550 and *β*^*^ = 0.24. The inset in each panel shows the predicted behaviors of *H*(*t*) and *I*(*t*) towards the end of the epidemic.

**Supplementary Figure S13.**
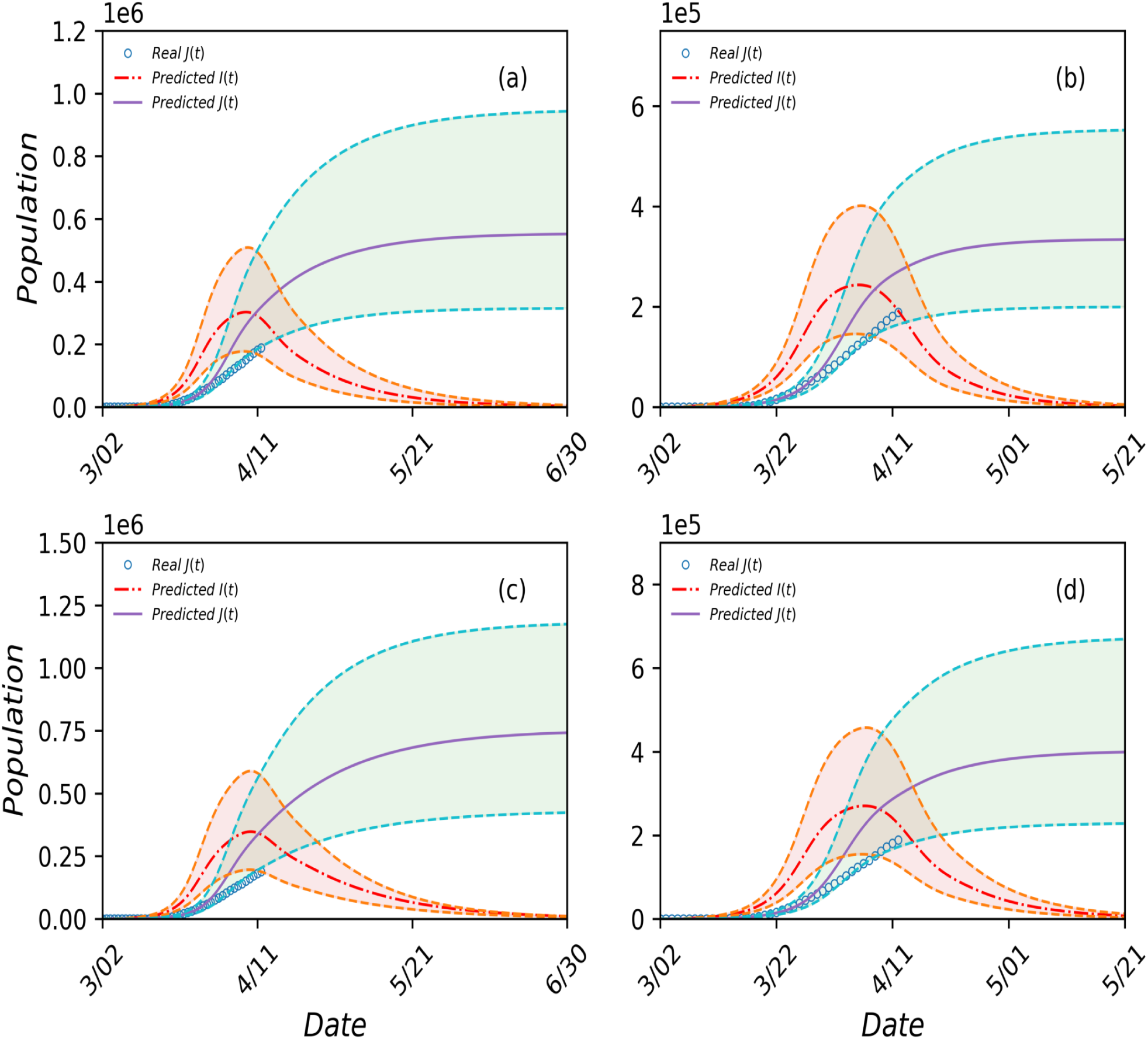
Uncertainty ranges of two representative quantities from the four predicted scenarios for the State of New York as an open system. The quantities are *J*(*t*) (light blue) and *I*(*t*) (light red), and the actual available daily data points of *J*(*t*) are denoted by the blue circles. The parameter settings are (a) *η* = 0.5 and λ= 0.18, (b) *η* = 0.5 and λ= 0.24, (c) *η* = 0.8 and λ= 0.18, and (d) *η* = 0.8 and λ= 0.24. For (a) and (b), optimization of the model equations gives *H*\*(0) = 3400 and *β*\* = 0.27, where the 95% confidence intervals for the former and latter are (2800, 4049) and (0.2565, 0.2840), respectively. For (c) and (d), the corresponding values are *H*\*(0) = 8600 and *β*\* = 0.25 with the respective 95% confidence interval (6897, 10465) and (0.2348, 0.2659).

**Supplementary Figure S14.**
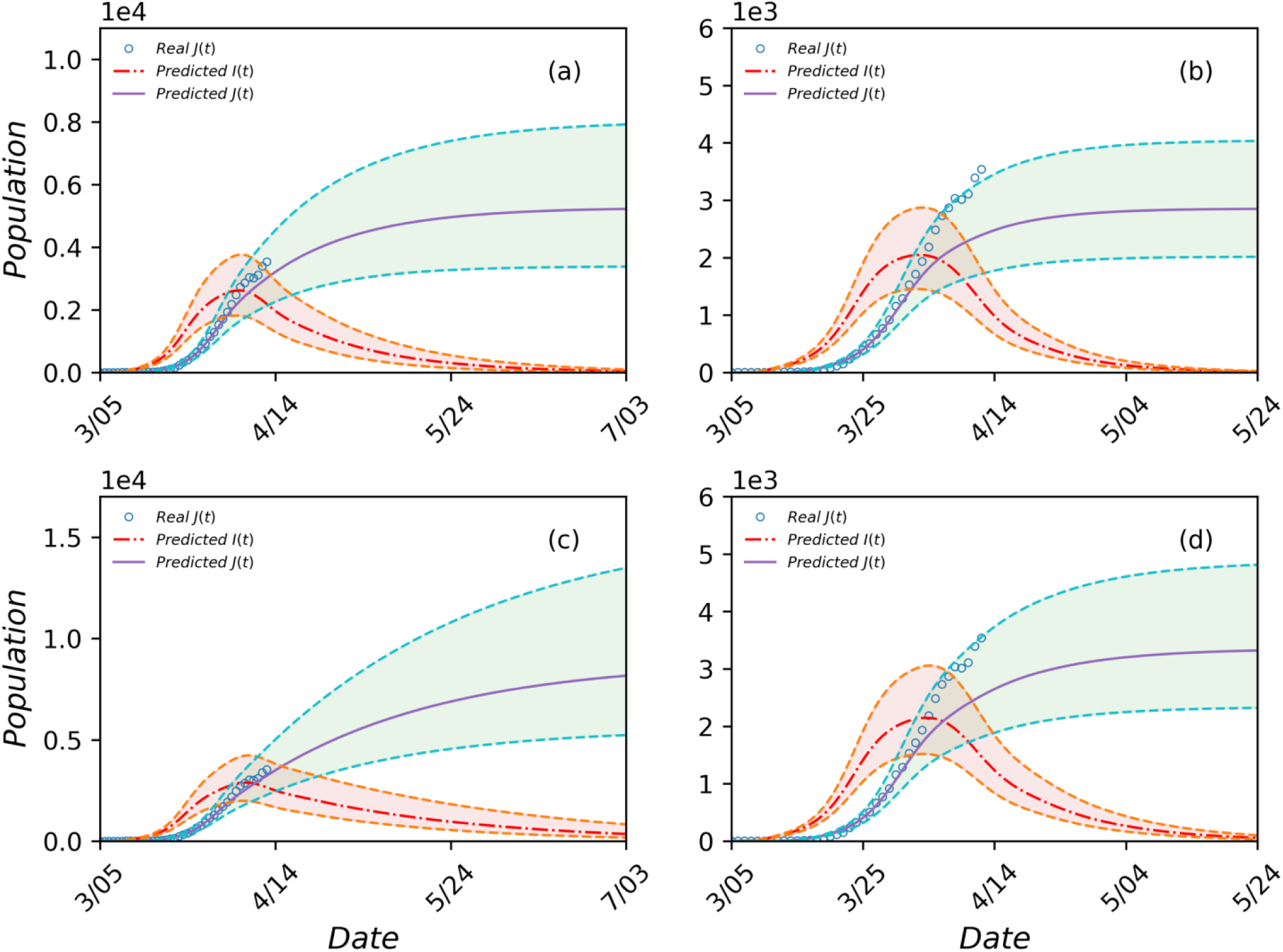
Uncertainty ranges of two representative quantities from the four predicted scenarios for the State of Arizona as an open system. The legends and settings are the same as those in Fig. S13. For (a) and (b), the optimally estimated parameter values are *H*\*(0) = 90 and *β*^*^ = 0.27 with the respective 95% confidence intervals (78, 103) and (0.2580, 0.2820). For (c) and (d), the corresponding values are *H*\*(0) = 260 and *β*^*^ = 0.24 with the respective 95% confidence interval (223, 293) and (0.2285, 0.2533).

**Supplementary Figure S15.**
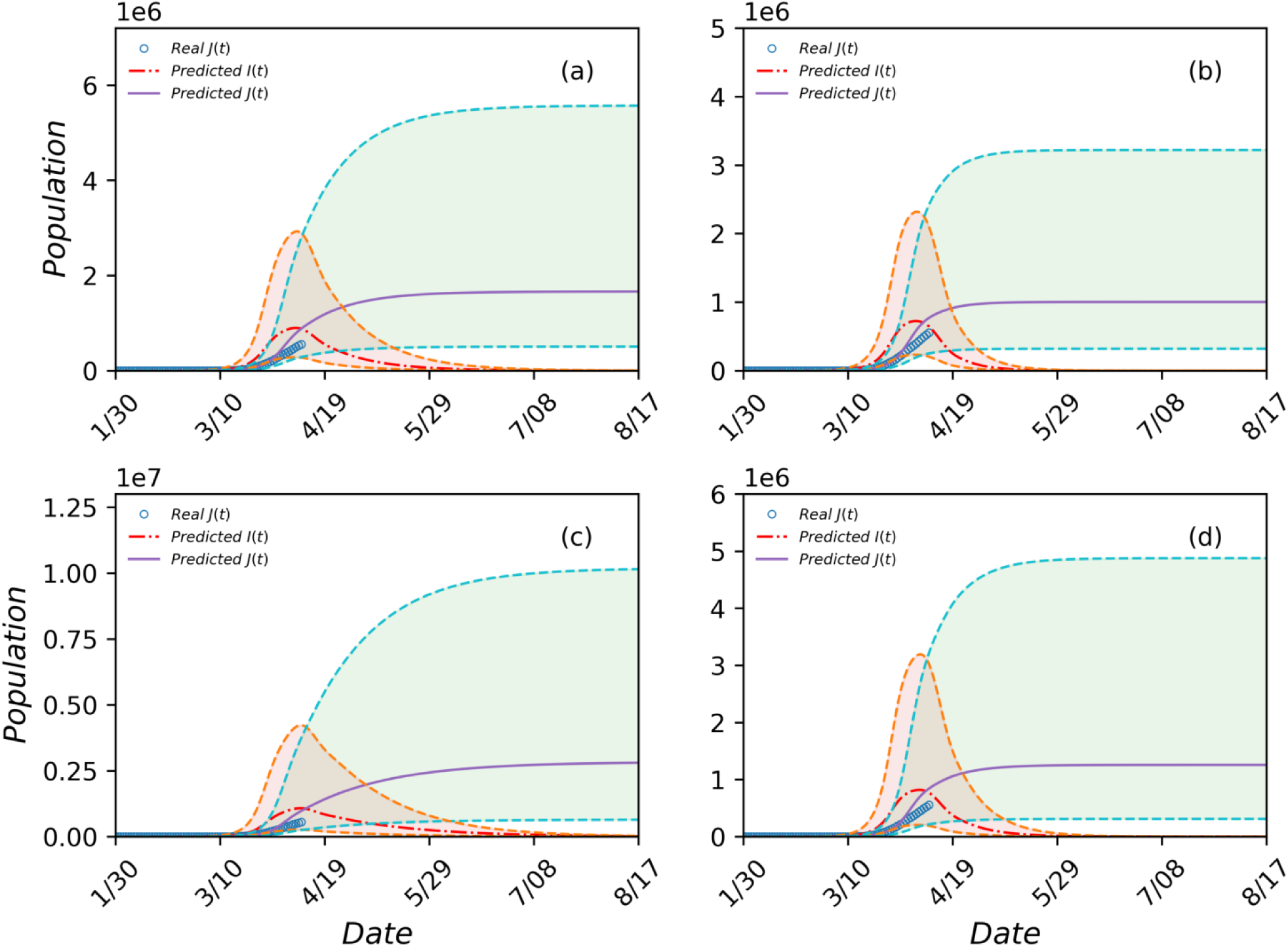
Uncertainty ranges of two representative quantities from the four predicted scenarios for the entire country of USA as a closed system. The legends and settings are the same as those in Fig. S13. For (a) and (b), the optimally estimated parameter values are *H*\*(0) = 20 and *β*^*^ = 0.26 with the respective 95% confidence intervals (12, 32) and (0.2498, 0.2715). For (c) and (d), the corresponding values are *H*\*(0) = 35 and *β*^*^ = 0.24 with the respective 95% confidence interval (18, 60) and (0.2281, 0.2543).

